# Phenotypes of disease severity in a cohort of hospitalized COVID-19 patients: results from the IMPACC study

**DOI:** 10.1101/2022.07.02.22273396

**Authors:** Al Ozonoff, Joanna Schaenman, Naresh Doni Jayavelu, Carly E. Milliren, Carolyn S. Calfee, Charles B. Cairns, Monica Kraft, Lindsey R. Baden, Albert C. Shaw, Florian Krammer, Harm van Bakel, Denise A. Esserman, Shanshan Liu, Ana Fernandez Sesma, Viviana Simon, David A. Hafler, Ruth R. Montgomery, Steven H. Kleinstein, Ofer Levy, Christian Bime, Elias K. Haddad, David J. Erle, Bali Pulendran, Kari C. Nadeau, Mark M Davis, Catherine L. Hough, William B. Messer, Nelson I Agudelo Higuita, Jordan P. Metcalf, Mark A. Atkinson, Scott C. Brakenridge, David Corry, Farrah Kheradmand, Lauren I. R. Ehrlich, Esther Melamed, Grace A. McComsey, Rafick Sekaly, Joann Diray-Arce, Bjoern Peters, Alison D. Augustine, Elaine F. Reed, Matthew C. Altman, Patrice M. Becker, Nadine Rouphael, the IMPACC study group members

**Author notes:** co-first authors (Al Ozonoff, Joanna Schaenman, Naresh Doni Jayavelu, Carly E. Milliren). co-senior authors (Matthew C. Altman, Patrice M. Becker, Nadine Rouphael).

## Abstract

**Background:** Better understanding of the association between characteristics of patients hospitalized with coronavirus disease 2019 (COVID-19) and outcome is needed to further improve upon patient management.

**Methods:** <u>Im</u>muno<u>p</u>henotyping <u>A</u>ssessment in a <u>C</u>OVID-19 <u>C</u>ohort (IMPACC) is a prospective, observational study of 1,164 patients from 20 hospitals across the United States. Disease severity was assessed using a 7-point ordinal scale based on degree of respiratory illness. Patients were prospectively surveyed for 1 year after discharge for post-acute sequalae of COVID-19 (PASC) through quarterly surveys. Demographics, comorbidities, radiographic findings, clinical laboratory values, SARS-CoV-2 PCR and serology were captured over a 28-day period. Multivariable logistic regression was performed.

**Findings:** The median age was 59 years (interquartile range [IQR] 20); 711 (61%) were men; overall mortality was 14%, and 228 (20%) required invasive mechanical ventilation. Unsupervised clustering of ordinal score over time revealed distinct disease course trajectories. Risk factors associated with prolonged hospitalization or death by day 28 included age ≥ 65 years (odds ratio [OR], 2.01; 95% CI 1.28-3.17), Hispanic ethnicity (OR, 1.71; 95% CI 1.13-2.57), elevated baseline creatinine (OR 2.80; 95% CI 1.63-4.80) or troponin (OR 1.89; 95% 1.03-3.47), baseline lymphopenia (OR 2.19; 95% CI 1.61-2.97), presence of infiltrate by chest imaging (OR 3.16; 95% CI 1.96-5.10), and high SARS-CoV2 viral load (OR 1.53; 95% CI 1.17-2.00). Fatal cases had the lowest ratio of SARS-CoV-2 antibody to viral load levels compared to other trajectories over time (p=0.001). 589 survivors (51%) completed at least one survey at follow-up with 305 (52%) having at least one symptom consistent with PASC, most commonly dyspnea (56% among symptomatic patients). Female sex was the only associated risk factor for PASC.

**Interpretation:** Integration of PCR cycle threshold, and antibody values with demographics, comorbidities, and laboratory/radiographic findings identified risk factors for 28-day outcome severity, though only female sex was associated with PASC. Longitudinal clinical phenotyping offers important insights, and provides a framework for immunophenotyping for acute and long COVID-19.

**Funding:** NIH

**RESEARCH IN CONTEXT:** *Evidence before this study:* We did a systematic search of the PubMed database from January 1^st^, 2020 until April 24^th^, 2022 using the search terms: “hospitalized” AND “SARS-CoV-2” OR “COVID-19” AND “Prospective” AND “Antibody” OR “PCR” OR “long term follow up” and applying the following filters: “Multicenter Study” AND “Observational Study”. No language restrictions were applied. While clinical, laboratory, and radiographic features associated with severe COVID-19 in hospitalized adults have been described, description of the kinetics of SARS-CoV-2 specific assays available to clinicians (e.g. PCR and binding antibody) and their integration with other variables is scarce for both short and long term follow up. The current literature is comprised of several studies with small sample size, cross-sectional design with laboratory data typically only recorded at a single point in time (e.g., on admission), limited clinical characteristics, variable duration of follow up, single-center setting, retrospective analyses, kinetics of either PCR or antibody testing but not both, and outcomes such as death or, mechanical ventilation that do not allow delineation of variations in clinical course.

*Added value of this study:* In our large longitudinal multicenter cohort, the description of outcome severity, was not limited to survival versus death, but encompassed a clinical trajectory approach leveraging longitudinal data based on time in hospital, disease severity by ordinal scale based on degree of respiratory illness, and presence or absence of limitations at discharge. Fatal COVID-19 cases had the lowest ratio of antibody to viral load levels over time as compared to non-fatal cases. Integration of PCR cycle threshold and antibody values with demographics, baseline comorbidities, and laboratory/radiographic findings identified additional risk factors for outcome severity over the first 28 days. However, female sex was the only variable associated with persistence of symptoms over time. Persistence of symptoms was not associated with clinical trajectory over the first 28 days, nor with antibody/viral loads from the acute phase.

*Implications of all the available evidence:* The described calculated ratio (binding IgG/PCR Ct value) is unique compared to other studies, reflecting host pathogen interactions and representing an accessible approach for patient risk stratification. Integration of SARS-CoV-2 viral load and binding antibody kinetics with other laboratory as well as clinical characteristics in hospitalized COVID-19 patients can identify patients likely to have the most severe short-term outcomes, but is not predictive of symptom persistence at one year post-discharge.

## INTRODUCTION

A comprehensive understanding of the interplay between pathogen, and host in modulating the disease course will enable identification of potential biomarkers, and host-directed therapeutic interventions against severe acute respiratory syndrome coronavirus 2 (SARS-CoV-2), and post-acute sequalae of coronavirus disease 2019 (COVID-19) (PASC). Mobilization of the global scientific community has produced significant findings with unprecedented speed. ^1^ However, these studies had limitations including small sample size, cross-sectional design with laboratory data typically only recorded at a single point in time (e.g., on admission), limited clinical characteristics, variable duration of follow up, single-center setting, retrospective analyses, and basic outcomes (e.g. death, mechanical ventilation). Integration of longitudinal data is an attractive approach to characterize disease severity, and better understand patient outcomes by encompassing the totality of the disease course in terms of patient complications, persistence of symptoms and resource utilization. In contrast to categorical analysis of outcomes at a prespecified timepoint, using a longitudinal approach provides insights into mechanisms underlying the diverse clinical course observed in a large group of geographically, and demographically diverse hospitalized patients with COVID-19.

The National Institute of Allergy and Infectious Diseases (NIAID), National Institutes of Health (NIH) prospective longitudinal cohort study, <u>Im</u>muno<u>P</u>henotyping <u>A</u>ssessment in a <u>C</u>OVID-19 <u>C</u>ohort (or IMPACC [NCT04378777]), enrolled symptomatic, molecularly confirmed hospitalized COVID-19 patients from 20 different hospitals across the US. IMPACC prospectively collected clinical, laboratory, and radiographic data along with longitudinal biologic sampling of blood, and respiratory secretions for in-depth immunologic, and virologic testing with a one year follow up post discharge.^2^

## METHODS

### Ethics

NIAID staff conferred with the Department of Health and Human Services Office for Human Research Protections (OHRP) regarding potential applicability of the public health surveillance exception [45CFR46.102(l)(2)] to the IMPACC study protocol. OHRP concurred that the study satisfied criteria for the public health surveillance exception, and the IMPACC study team sent the study protocol, and participant information sheet for review, and assessment to institutional review boards (IRBs) at participating institutions. Twelve institutions elected to conduct the study as public health surveillance, while 3 sites with prior IRB-approved biobanking protocols elected to integrate and conduct IMPACC under their institutional protocols (University of Texas at Austin, IRB 2020-04-0117; University of California San Francisco, IRB 20-30497; Case Western reserve university, IRB STUDY20200573) with informed consent requirements. Participants enrolled under the public health surveillance exclusion were provided information sheets describing the study, samples to be collected, and plans for data de-identification, and use. Those that requested not to participate after reviewing the information sheet were not enrolled. In addition, participants did not receive compensation for study participation while inpatient, and subsequently were offered compensation during outpatient follow-ups.

### Study Design and Setting

The study followed the Strengthening the Reporting of Observational Studies in Epidemiology (STROBE) guidelines for reporting observational studies.^3^ The design of the IMPACC study has been previously published^2^ (see online supplement).

### Study Participants

Patients 18 years and older admitted to 20 US hospitals (affiliated with 15 academic institutions) were enrolled within 48 hours of hospital admission. Only symptomatic cases with confirmed positive SARS-CoV-2 PCR were followed longitudinally.

### Data Collection, Study Variables, and Biologic Samples

Specific data elements were acquired via review of electronic medical records: participant demographic characteristics, and comorbidities, presenting signs, or symptoms, and onset, medications, diagnostic investigations (predefined laboratory values, and radiographic findings), and relevant clinical outcomes (oxygen- and ventilatory-support requirement, medications used, complications). Biologic samples consisted of blood, and mid-turbinate nasal swabs (staff or self collected; with step-by-step instructions provided to both staff and study participants). The timepoints were as follows: enrollment (day 1), and days 4, 7, 14, 21, and 28 post hospital admission (and if feasible, in discharged patients, days 14, and 28). Data lock on the data collected through day 28 from admission was performed on November 11^th^, 2021. Selected information was available after discharge for self-reported vaccination status, recurrent SARS-CoV2 infection, and persistence of symptoms, as well as mortality in patients with at least one follow-up at 3, 6, 9 and/or 12 months after discharge. Data lock on the survey data was performed on April 7, 2022. The full study data collection forms are provided in the online supplement and deidentified data is available upon request.

### Outcomes

Clinical severity of illness was assessed using a 7-point ordinal scale (OS) adapted from the World Health Organization COVID-19, and NIAID disease ordinal severity scales,^4^ (OS1= Not hospitalized, no limitations; OS2= Not hospitalized, activity limitations, or requires home O_2_; OS3= Hospitalized, not requiring supplemental O_2_; OS4= Hospitalized, requiring O_2_; OS5= Hospitalized on non-invasive ventilation, or high-flow O_2_; OS6= Hospitalized on invasive mechanical ventilation, and/or extracorporeal membrane oxygenation (ECMO); OS7=Death). The 7-point OS for respiratory status was calculated at each inpatient timepoint. Length of hospital stay, complications, and other protocol-defined outcomes were assessed over 28 days and persistence of symptoms, reinfections as well as 28 day and delayed mortality were assessed for the duration of the study.

### SARS-CoV-2 PCR

SARS-CoV-2 viral load was assessed by a central laboratory from nasal swab samples at each time point by RT-PCR of the viral N1, and N2 genes (see online supplement)^5^.

### Viral Sequencing

Viral sequencing was performed by a central laboratory from one or more nasal swab samples per patient, ranked by lowest Ct value, to obtain at least one complete genome per patient. This yielded a total of 660 available genomes from 453 patients (see online supplement)^6^.

### Serology

At the time of publication, anti-SARS-CoV-2 spike (S), and receptor binding domain (RBD) antibodies, quantitated by enzyme-linked immunosorbent assay (ELISA) in serum,^7^ were available for the first 891 participants (see online supplement).

### Statistics

Longitudinal measures of ordinal scale over time were clustered using group-based trajectory modeling, a likelihood-based approach commonly used to group time series of clinical data. These models were implemented in an unsupervised fashion using SAS PROC TRAJ, using either 3, 4, 5, or 6 groups. For each specification of group number, polynomial curves up to cubic degree were allowed for each group. All models included timing of symptom onset relative to hospital admission as fixed effect. The best-fitting model, with respect to number of clusters and polynomial order, was selected using Akaike Information Criteria (AIC). A detailed description of the model implementation is provided in the online supplement.

Demographic, and clinical characteristics are reported as mean (standard deviation [SD]), median (interquartile range [IQR]), or frequency (percentage) compared across clinical trajectory groups using one-way ANOVA, Kruskal-Wallis tests, or chi-square tests as appropriate. Where distributions were skewed, continuous laboratory values were log-transformed prior to parametric analysis. Abnormal baseline laboratory values were defined based on site-specific normal range. Missing data were not imputed; however, labs that were not drawn were assumed to be normal. The extent of missing data is noted where applicable.

A series of multivariable logistic regression models was fit to identify demographic, and clinical risk factors associated with disease severity. All models included random intercept by enrollment site, and covariates were selected through a multi-step selection process (see Supplement). The covariates selected for the final model and included as fixed effects (see Figure 4) are: age, sex, race, Hispanic ethnicity, diabetes, chronic respiratory disease, presence of infiltrate on chest x-ray, lymphocyte count, creatinine level, troponin level, and N1 CT value. Ordinal, and logistic regressions identified laboratory variables associated with the overall ordinal clinical trajectory trend, and discriminative of two different trajectory groups at baseline. A mixed-effect generalized additive model identified variables whose 28 day longitudinal values differ across clinical trajectory groups. This model included fixed effects for age and sex, and random effects for enrollment site and participant. Further details of the model specifications, and approaches for model selection are provided in the online supplement.

All analyses were performed in SAS (v9.4, Cary, NC), and R (v4.1, XXX) using an alpha-level of 0.05.

### Role of the funders

The study was funded by project grants and cooperative agreements awarded by the National Institute of Allergy and Infectious Disease (NIAID). NIAID project scientists participated collaboratively in study design, data analyses, interpretation, and writing of the report but not in data collection. The manuscript content is solely the responsibility of the authors and does not necessarily represent the official views of the National Institute of Allergy and Infectious Diseases, the National Institutes of Health or any other Agency of the United States Government.

The authors are solely responsible for the study design, data collection, interpretation, manuscript preparation, and decision to submit the manuscript.

## RESULTS

Between May 5^th^, 2020 and March 19^th^, 2021, 1,286 patients were enrolled across 20 US hospitals; 1,243 met eligibility criteria, and 1,164 were included in the final analysis cohort with 589 being included in the post-hospitalization survey cohort (FIGURE 1). Clinical characteristics, baseline radiographic findings, and laboratory testing of the study cohort are provided in TABLE 1. The median age was 59 years (IQR 20), and 711 (61%) were men. 259 (22%) were Black/African American, and 363 (31%) were Hispanic/Latinx. 1,096 (94%) had at least one comorbidity, including hypertension (677; 58%), diabetes (427; 37%), chronic respiratory disease (234; 20%), asthma (174; 15%), chronic cardiac disease (315; 27%), and chronic kidney disease (178; 15%). Current, or prior history of smoking, or vaping was reported in 377 (32%) patients. 58 (5%) were organ transplant recipients, and 21 (2%) were patients living with HIV. The median body mass index (BMI) was 31.3 kg/m^2^ (IQR 9.6). Most patients (872; 75%) presented within 2 weeks of onset of symptoms. 276 (24%) were admitted directly to the intensive care unit (ICU) with 137 (12%) initially requiring mechanical ventilation, or ECMO. Almost half of the cohort presented with elevated D-dimer (>0.5 mg/L) (596; 51%), and C-reactive protein (CRP) (≥10 mg/L) (499; 43%). 139 (12%) patients had severe lymphopenia (<500 cells/microliter), and 59 (5%) had thrombocytopenia (<100,000 cells/microliter) at enrollment. A subset presented with evidence of renal insufficiency (creatinine ≥1.5 mg/dL; 186; 16%), liver dysfunction (alanine transaminase (ALT) ≥1.5 site specific upper limit of normal; 203; 17%), and/or myocardial injury (95 out of 445 available values with troponin >0.4 ng/mL; 8%). Most patients had opacities consistent with pneumonia on chest imaging on enrollment (800; 72%). The median SOFA (Sequential Organ Failure Assessment) score at baseline was 1 (IQR: 2).

**FIGURE 1.**
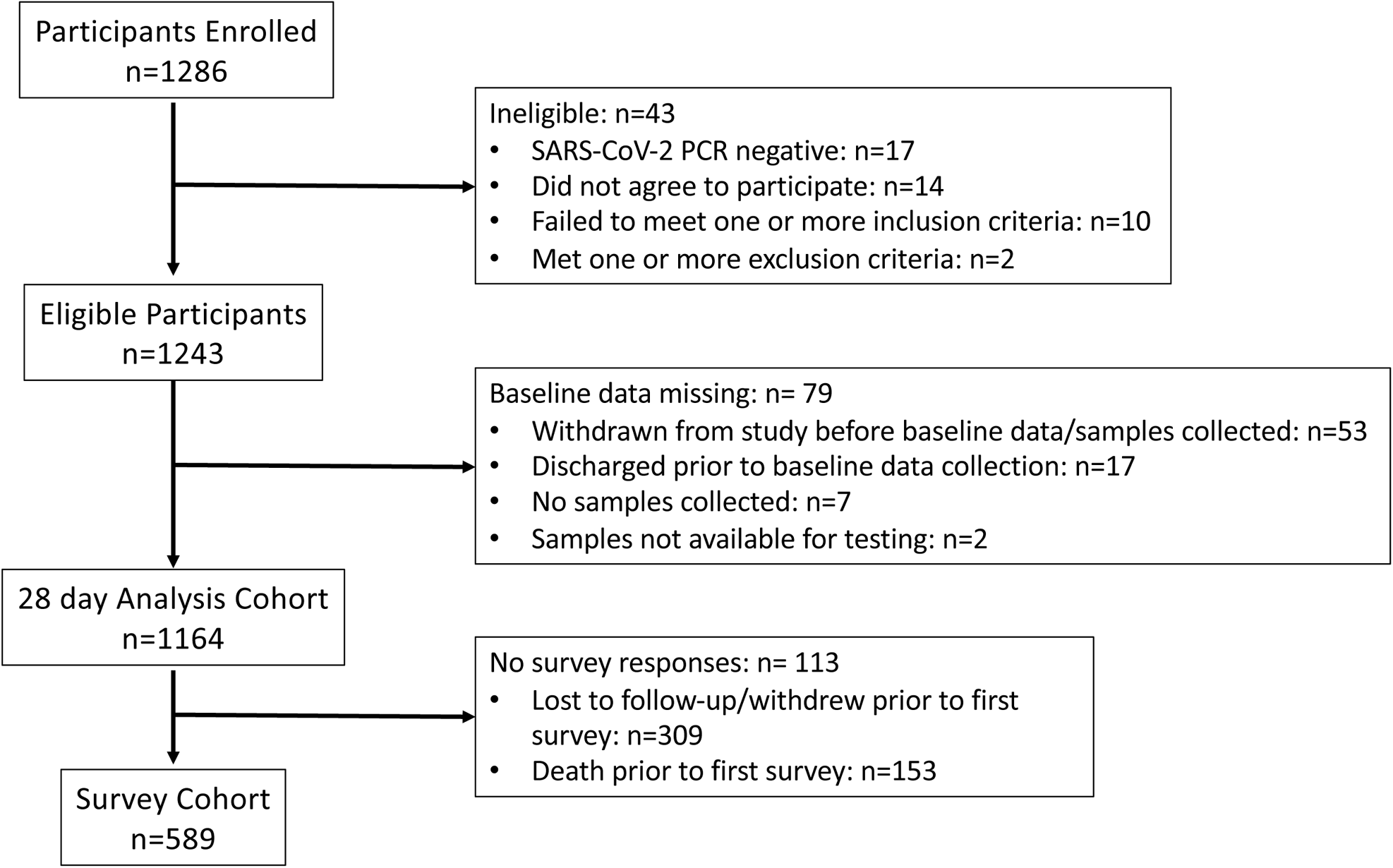
Strengthening the Reporting of Observational Studies in Epidemiology (STROBE) Cohort Diagram

**TABLE 1.**
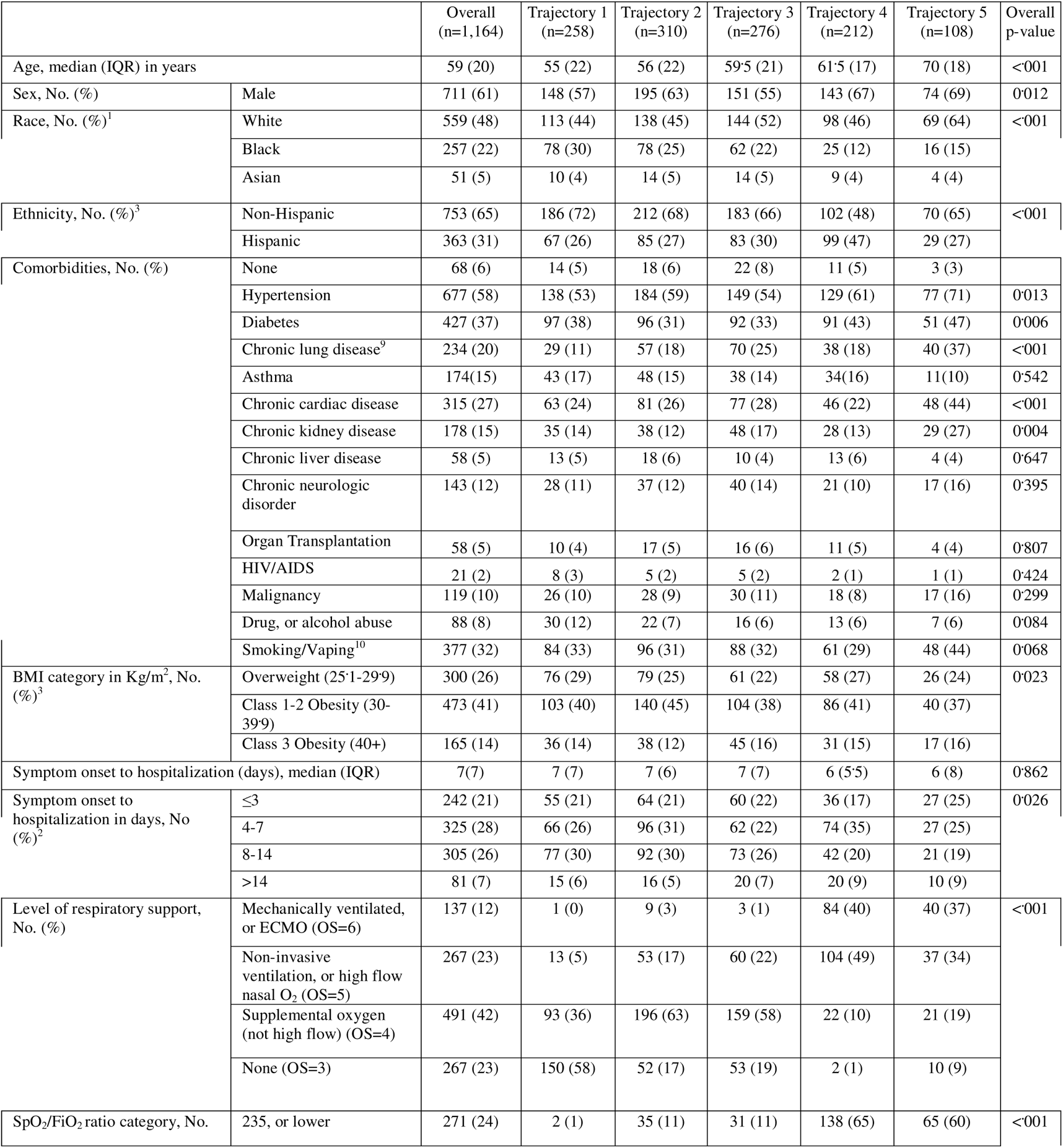
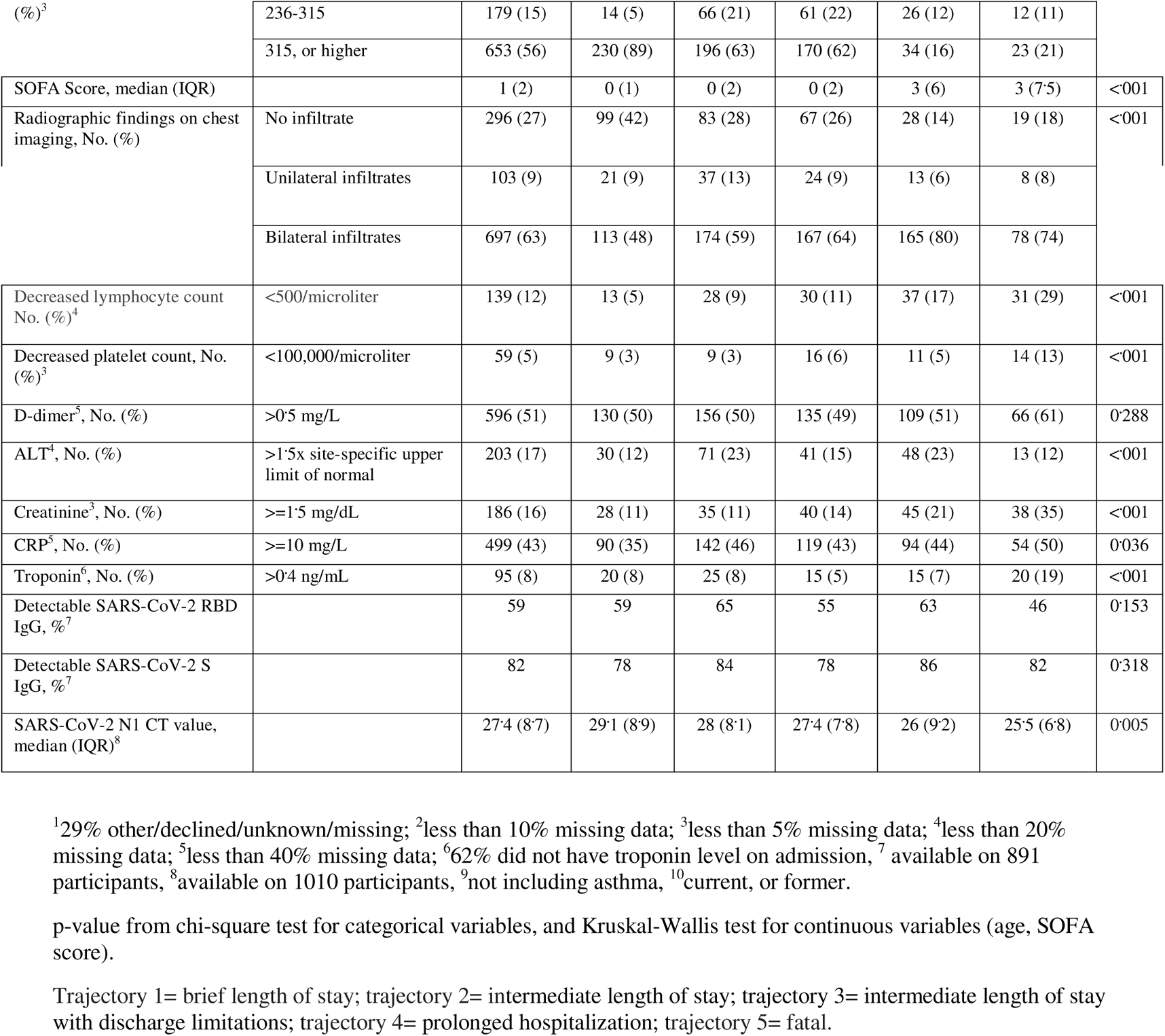
Demographics, Clinical Characteristics, Comorbidities, Radiographic Findings, and Laboratory Testing of Cohort Participants at Baseline (N=1,164)

The median length of hospital stay was 6 days (IQR 7). Twenty-eight day mortality was 9%, and did not differ across the duration of the study (divided into quarters; p=0.84) (FIGURE 1 SUPPLEMENT); the overall mortality 12 months post hospital discharge was 14%. At day 28, 70 (6%) remained hospitalized. During their hospitalization, 228 (20%) required invasive mechanical ventilation, 176 (15%) experienced ICU level care escalation, and 958 (82%) had at least one pre-specified complication (TABLE 1 SUPPLEMENT) including acute renal injury/failure (249; 21%), shock requiring use of pressors (173; 15%), bacteremia (113; 10%). 791 (68%) received systemic steroids (786; 68%), and 725 (62%) received remdesivir (TABLE 2 SUPPLEMENT).

To capture the dynamics of clinical course of disease, we analyzed patient characteristics through an unsupervised clustering of respiratory OS over time. Five disease course trajectories were identified (FIGURE 2): brief length of stay (trajectory 1: n=258; 22%); intermediate length of stay (trajectory 2: n=310; 27%); intermediate length of stay with discharge limitations (trajectory 3: n=276; 24%); prolonged hospitalization (trajectory 4: n=212;18%); and fatal (trajectory 5: n= 108; 9%).

**FIGURE 2:**
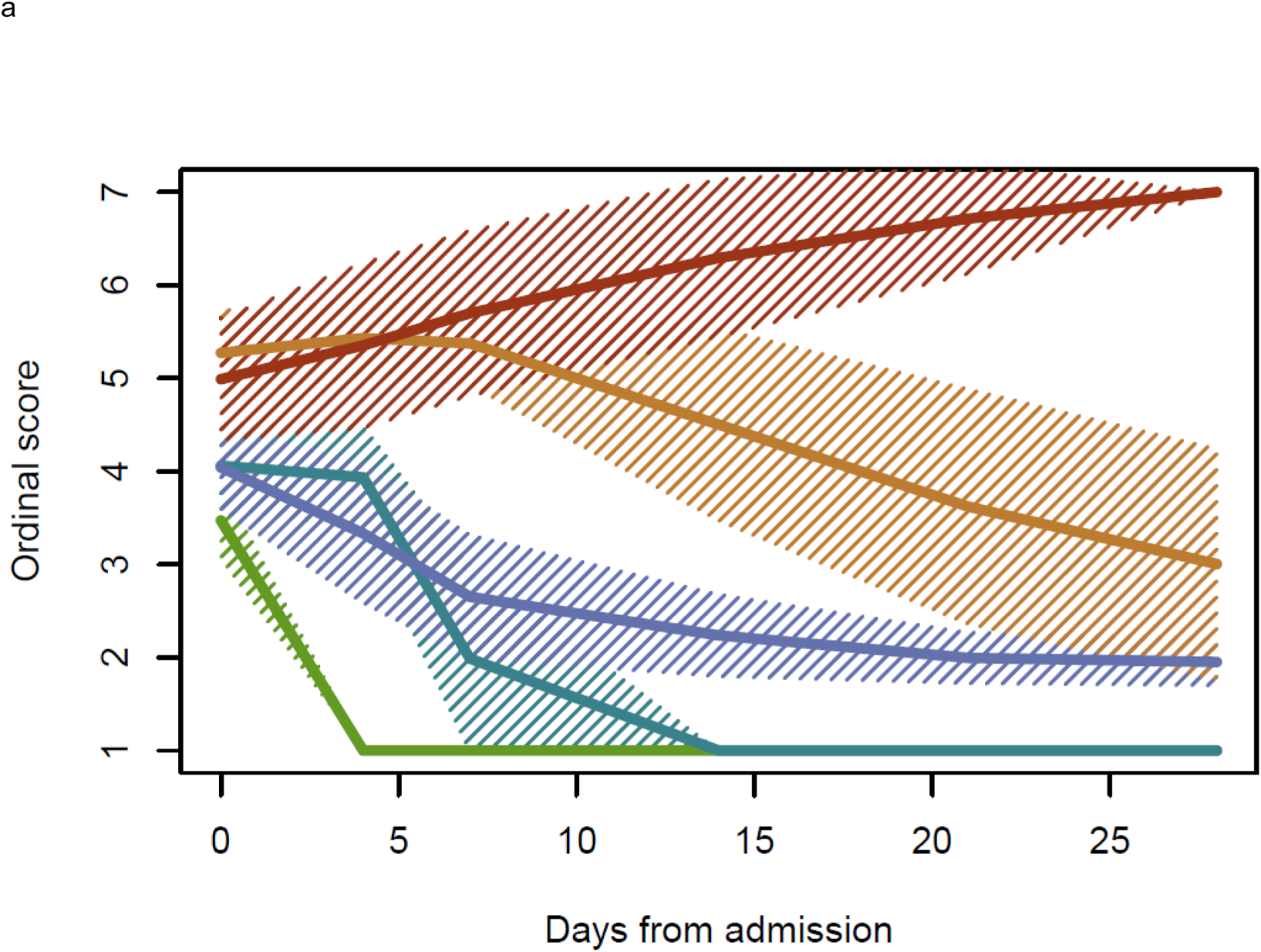
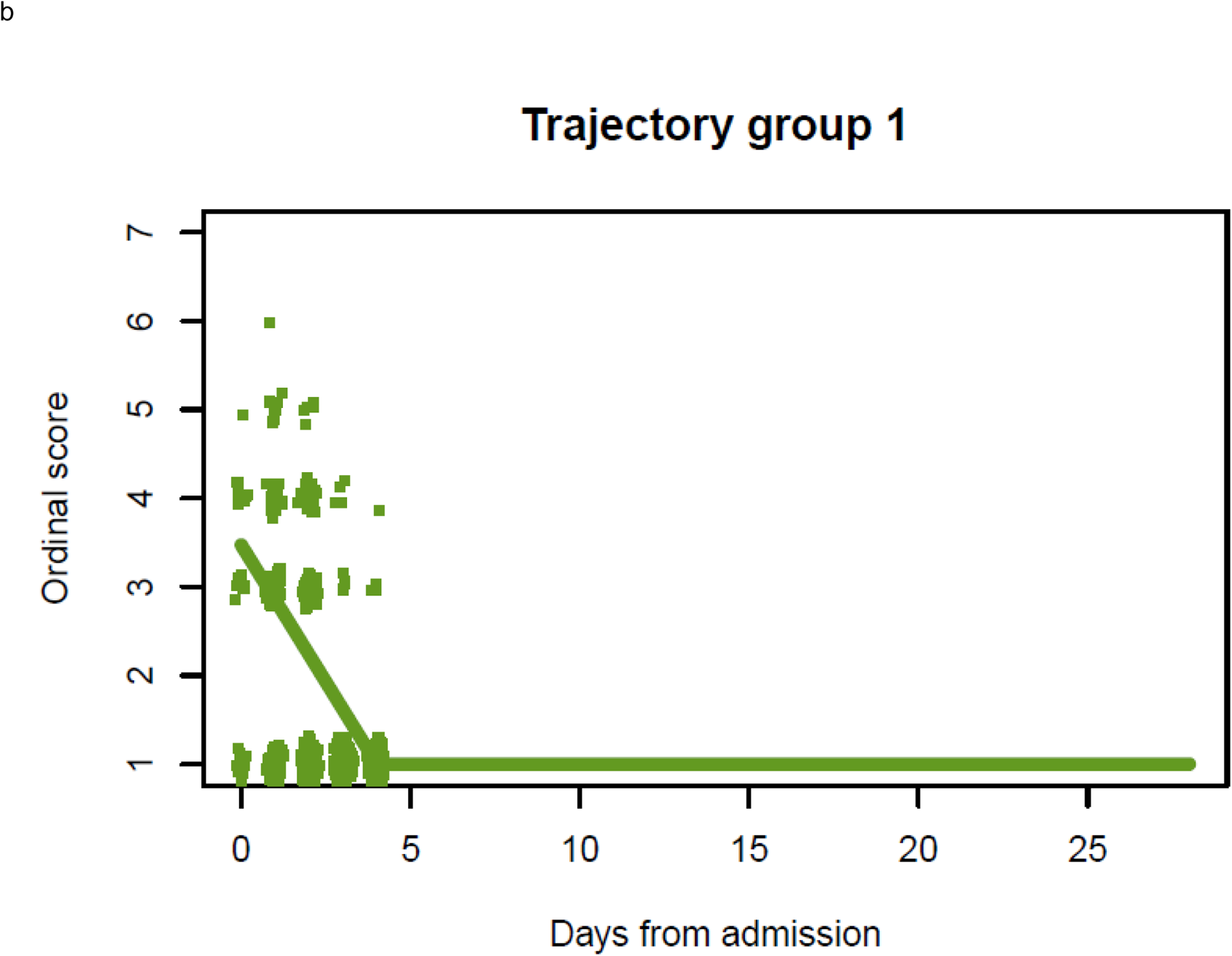
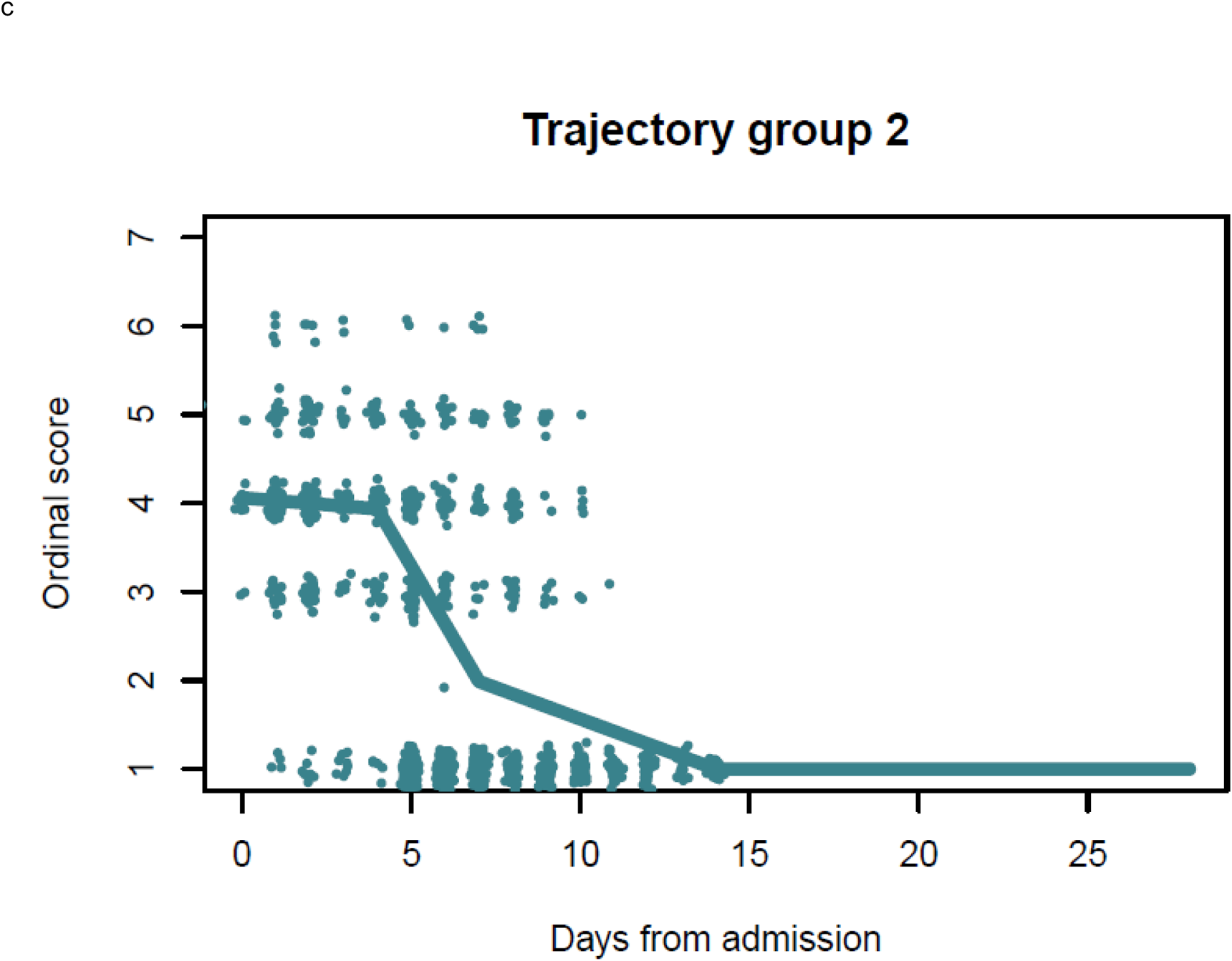
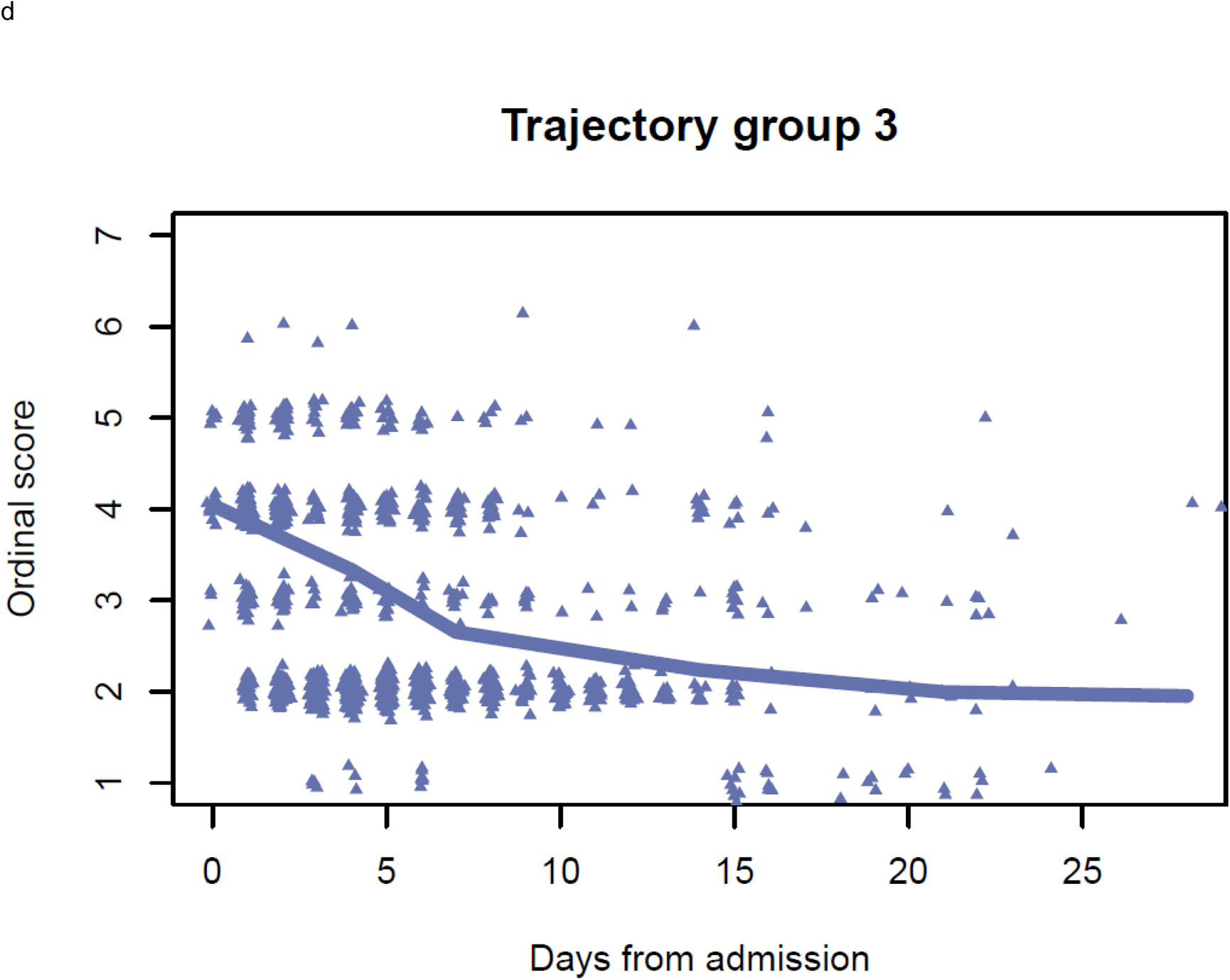
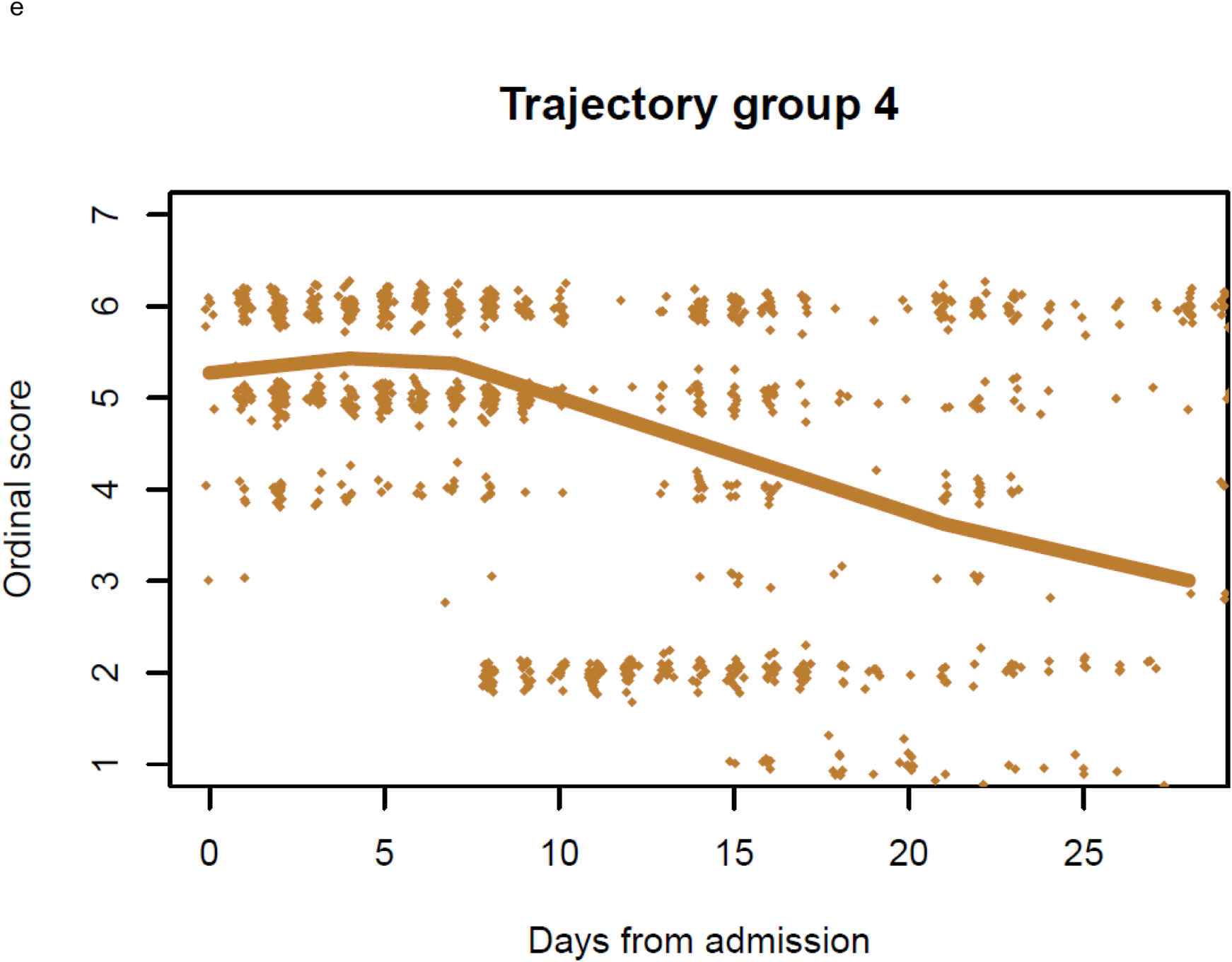
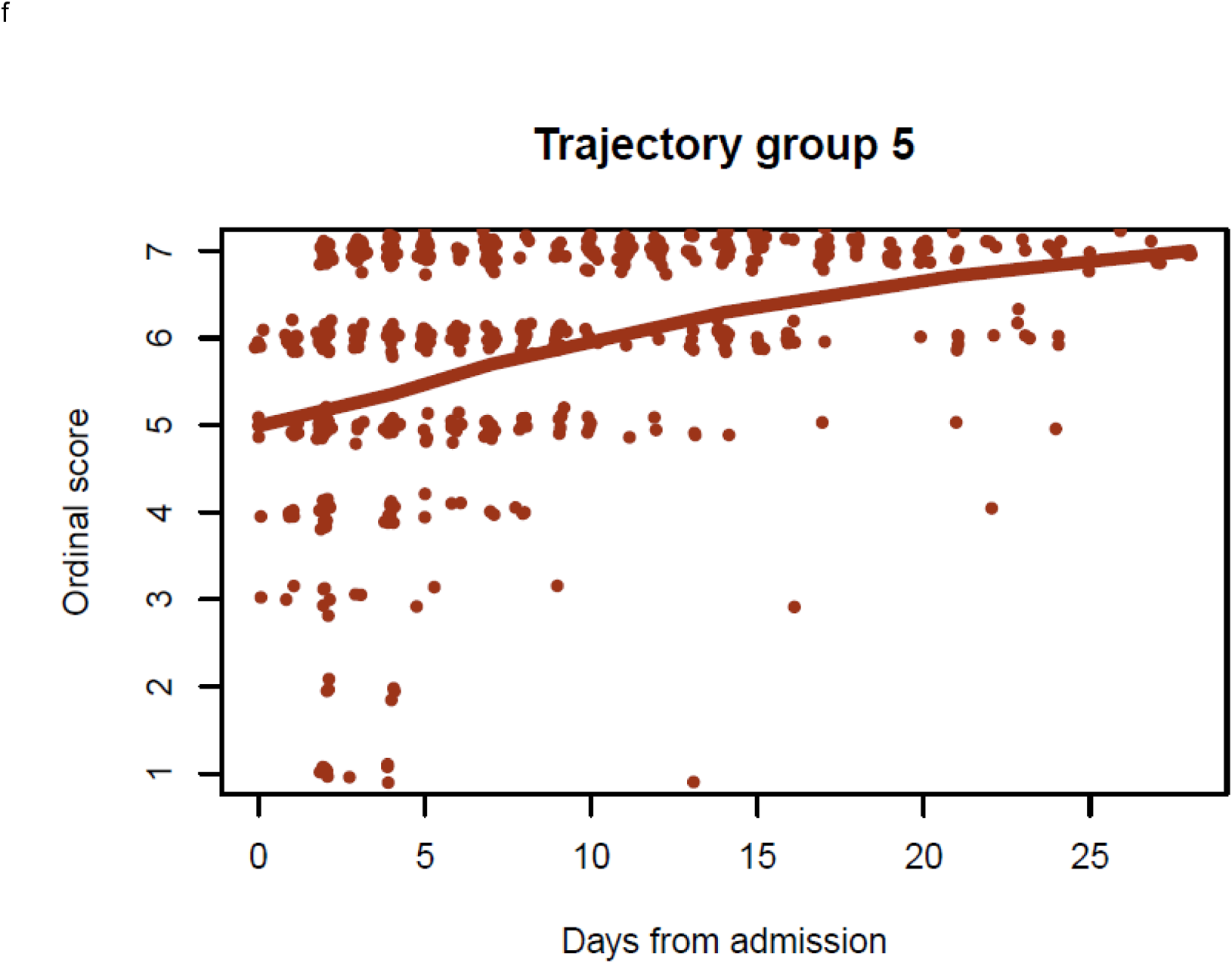
Classification of clinical trajectories. Longitudinal trajectories of clinical respiratory status were classified into one of five groups. Mean ordinal score for each group are shown with IQR determined by normal approximation (i.e. group-specific mean +/- 0.674 SD) (panel a), estimated at each of the six visit timepoints as determined by study protocol. Separate group-specific mean plots are shown (panels b-f) with points representing observed participant-level (jittered) ordinal scores by days from admission. Trajectory 1= brief length of stay (green); trajectory 2= intermediate length of stay (teal); trajectory 3= intermediate length of stay with discharge limitations (blue); trajectory 4= prolonged hospitalization (orange); trajectory 5= fatal (red). Ordinal scores: OS1= Not hospitalized, no limitations; OS2= Not hospitalized, activity limitations, or requires home O_2_; OS3= Hospitalized, not requiring supplemental O_2_; OS4= Hospitalized, requiring O_2_; OS5= Hospitalized on non-invasive ventilation, or high-flow O_2_; OS6= Hospitalized on invasive mechanical ventilation, and/or ECMO; OS7=Death.

Selected demographic characteristics, key comorbidities, radiographic, and laboratory findings were significantly associated with trajectory groups by univariate analysis (TABLE 1). In terms of symptoms at presentation, shortness of breath (p<0.001), and altered mental status (p<0.001) were associated with more severe disease, while gastrointestinal symptoms were associated with milder trajectories (p<0.001). The interval between onset of symptoms and hospitalization was not consistently associated with worsening outcome (Table 1). Higher markers of inflammation (i.e., CRP, lactate dehydrogenase, ferritin) differed by trajectories, but did not differ significantly between trajectories 4, and 5. An increased number of complications was noted with trajectories associated with higher OS (TABLE 1 SUPPLEMENT).

N1 SARS-CoV-2 PCR median cycle threshold (Ct) values at enrollment were significantly lower (indicating higher viral RNA levels) in patients with a more severe disease course (FIGURE 3a) with progressively lower Ct values from trajectory 1 through 5 (p=0.0053). This pattern persisted throughout the 28 day-period; namely, more severe trajectories maintained higher viral RNA levels (FIGURE 3b) (p<0.001). In the majority of trajectories, we observed a steady decline in viral RNA levels in the first week after hospitalization. However, uniquely in trajectory 5, the rate of decline was attenuated between the first, and second week of hospitalization, and plateaued at a Ct value below 30 during the 2^nd^ week of hospitalization (p=0.009), indicating persistence of viral RNA. In the other 4 trajectories, viral RNA levels continued to decline between 14, and 28 days after hospitalization. Viral RNA levels followed a similar pattern when analyzed by days from symptom onset S(FIGURE 4 SUPPLEMENT). Similar observations were also seen with N2 SARS-CoV-2 PCR (FIGURE 2 SUPPLEMENT, FIGURE 4 SUPPLEMENT). Adjustment for administration of remdesivir, or systemic glucocorticoids during the hospital course did not alter these findings (FIGURE 3 SUPPLEMENT). Viral sequencing, available in less than half of the cohort, revealed few variants of concern or interest that circulated during the study conduct [7 patients with alpha (B.1.1.7), 12 with epsilon (B.1.427 and B.1.429) and 4 with iota (B1.429)] (FIGURE 5 SUPPLEMENT).

**FIGURE 3.**
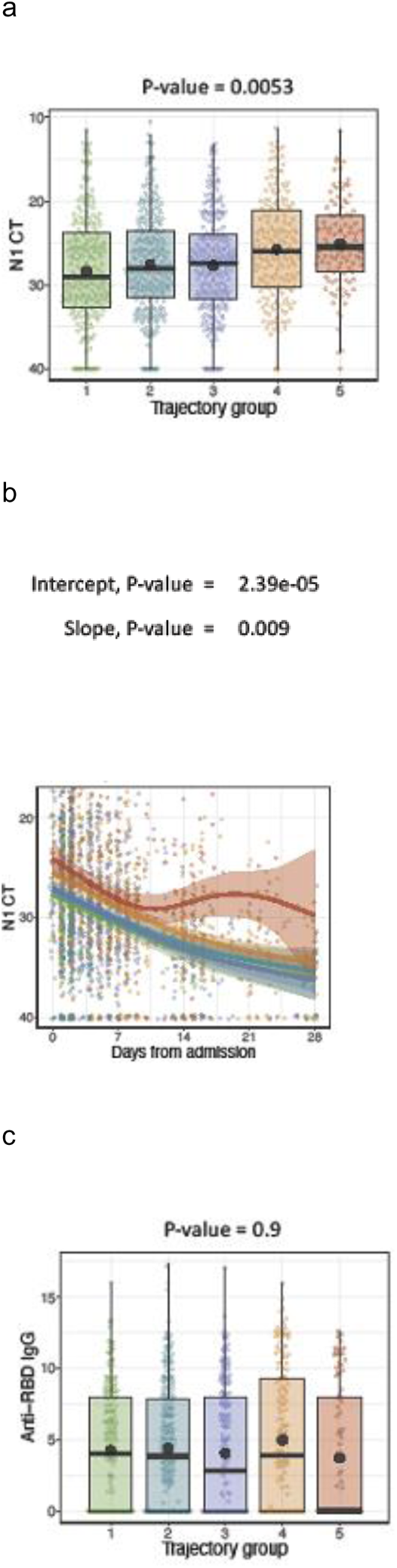
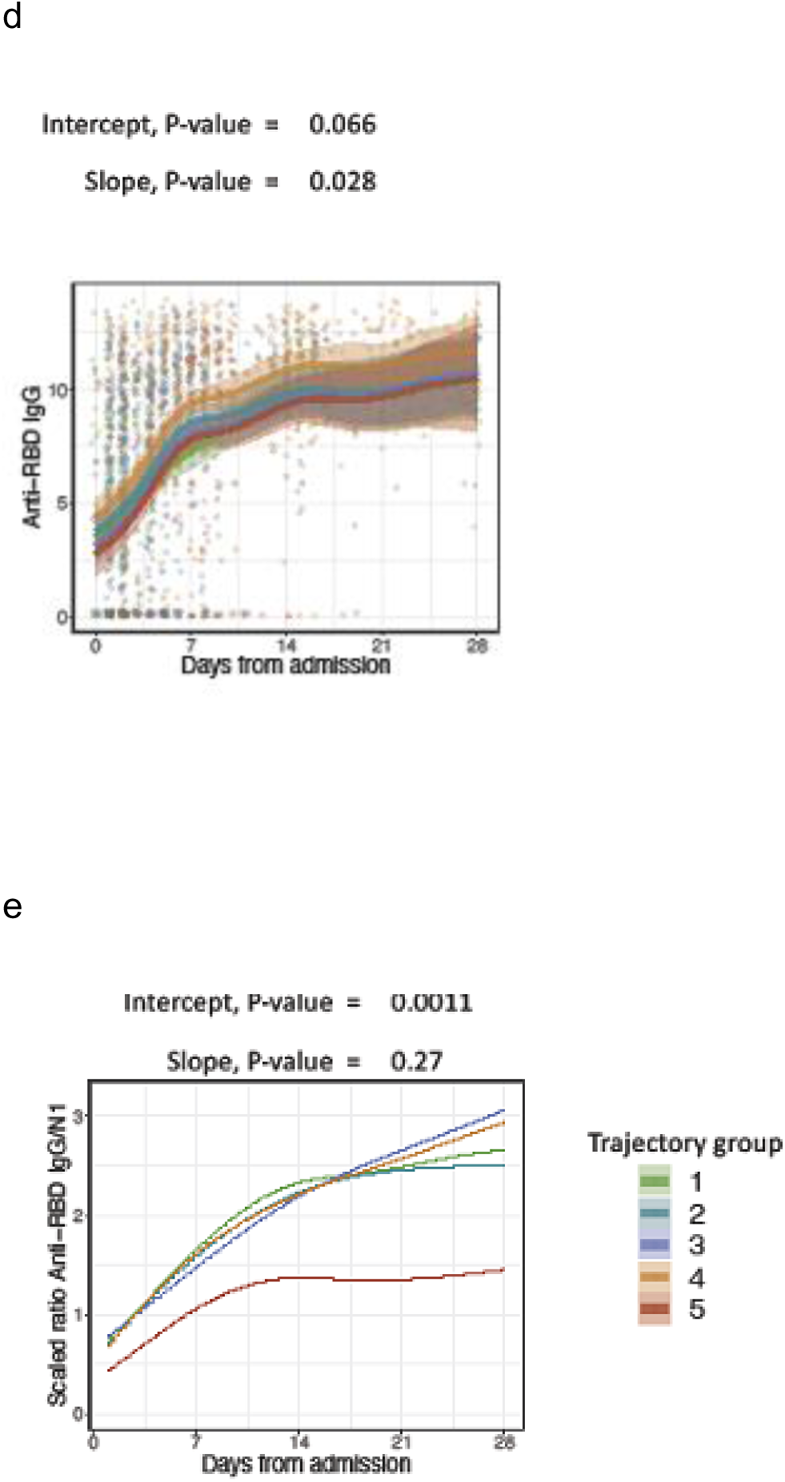
(a) N1 Ct baseline: Shown are the SARS-CoV-2 N1 gene Ct values during the first study collection timepoint which occurred at any time from admission until up to approximately 48 hours after admission. Shown are median values (horizontal lines), mean values (black points), interquartile ranges (boxes), and 1.5 IQR (whiskers), as well as all individual points. (b) N1 Ct over time: Shown are the SARS-CoV-2 N1 gene Ct values collected during the first 28 days of hospital admission. Trend lines for each trajectory represent the fit of a generalized additive model (GAM) to capture non-linear trends in the data. (c) RBD IgG baseline: Shown are the anti-RBD IgG values during the first study collection timepoint which occurred at any time from admission until up to approximately 48 hours after admission. Shown are median values (horizontal lines), mean values (black points), interquartile ranges (boxes), and 1.5 IQR (whiskers), as well as all individual points. (d) RBD IgG over time: Shown are the RBD IgG values collected during the first 28 days of hospital admission. Trend lines for each trajectory represent the fit of a generalized additive model (GAM) to capture non-linear trends in the data. (e) Ratio anti-RBD IgG/N1 Ct over time: Shown are the trends of the scaled ratio of anti-RBD IgG values divided by scaled SARS-CoV-2 N1 gene Ct values for time points collected during the first 28 days of hospital admission, emphasizing the divergence of trajectory 5 from the other 5 trajectories in this measure. Trajectory 1= brief length of stay (green); trajectory 2= intermediate length of stay (teal); trajectory 3= intermediate length of stay with discharge limitations (blue); trajectory 4= prolonged hospitalization (orange); trajectory 5= fatal (red).

Anti-S, and RBD IgG antibodies (FIGURE 3c, d, and FIGURE 2 SUPPLEMENT) were detectable on admission in 81.6%, and 58.8% of patients, respectively (TABLE 1). As expected, antibody levels were higher in patients who presented more than 7 days after symptom onset compared to those who presented within 7 days of symptom onset (FIGURE 4 SUPPLEMENT). In the subset of individuals presenting more than 7 days after symptom onset, significantly lower levels of anti-RBD antibody were observed in trajectory 5 compared to other trajectories (p=0.03-0.04). We observed a rapid increase in anti-RBD, and anti-S IgG levels in the first 7 days after admission followed by a moderate rise between 7, and 21 days, reaching a plateau towards the end of the 28-day period.

Analysis of the relationship between antibody levels and antibody to viral load ratio revealed associations with clinical trajectories. Throughout this 28-day window period, trajectory 4 participants had relatively higher anti-S, and RBD antibody levels compared to the other trajectories, whereas the lowest levels tended to be observed in trajectory 5 (p=0.066). Anti-RBD, and S IgG levels were similar, and correlated well (Pearson correlation, R=0.91, p<0.001). We calculated the scaled ratio of anti-RBD antibody levels to Ct values, and observed that trajectory 5 had significantly lower values of this ratio compared to the other four trajectories over the 28-day window period (p = 0.001) (FIGURE 3e).

Multivariable analyses comparing patients with short or intermediate length of stay (trajectories 1 to 3) to patients who had prolonged hospitalization or died (trajectories 4, and 5), suggested that those with more severe illness were more likely to be 65 years, or older (odds ratio [OR], 2.01; 95% CI 1.28-3.17), of Latinx ethnicity (OR, 1.71; 95% CI 1.13-2.57), had elevated baseline creatinine (OR 2.80; 95% CI 1.63-4.80), or troponin (OR 1.89; 95% 1.03-3.47), baseline lymphopenia (OR 2.19; 95% CI 1.61-2.97), presence of infiltrate by chest imaging (OR 3.16; 95% CI 1.96-5.10), and high SARS-CoV-2 viral burden (Ct < 27.4 (median value) (OR 1.53; 95% CI 1.17-2.00) (FIGURE 4). In multivariate analyses comparing patients who died (trajectory 5) with those with a prolonged hospitalization (trajectory 4), death was independently associated with older age (OR 1.81; 95% CI 1.24-2.64), chronic respiratory disease (OR 2.11; 95% 1.34-3.34), chronic kidney disease (OR 2.65; 95% CI 1.63-4.33), and low baseline platelet count (OR 2.68; 95% CI 1.13-6.36) (FIGURE 4).

**FIGURE 4.**
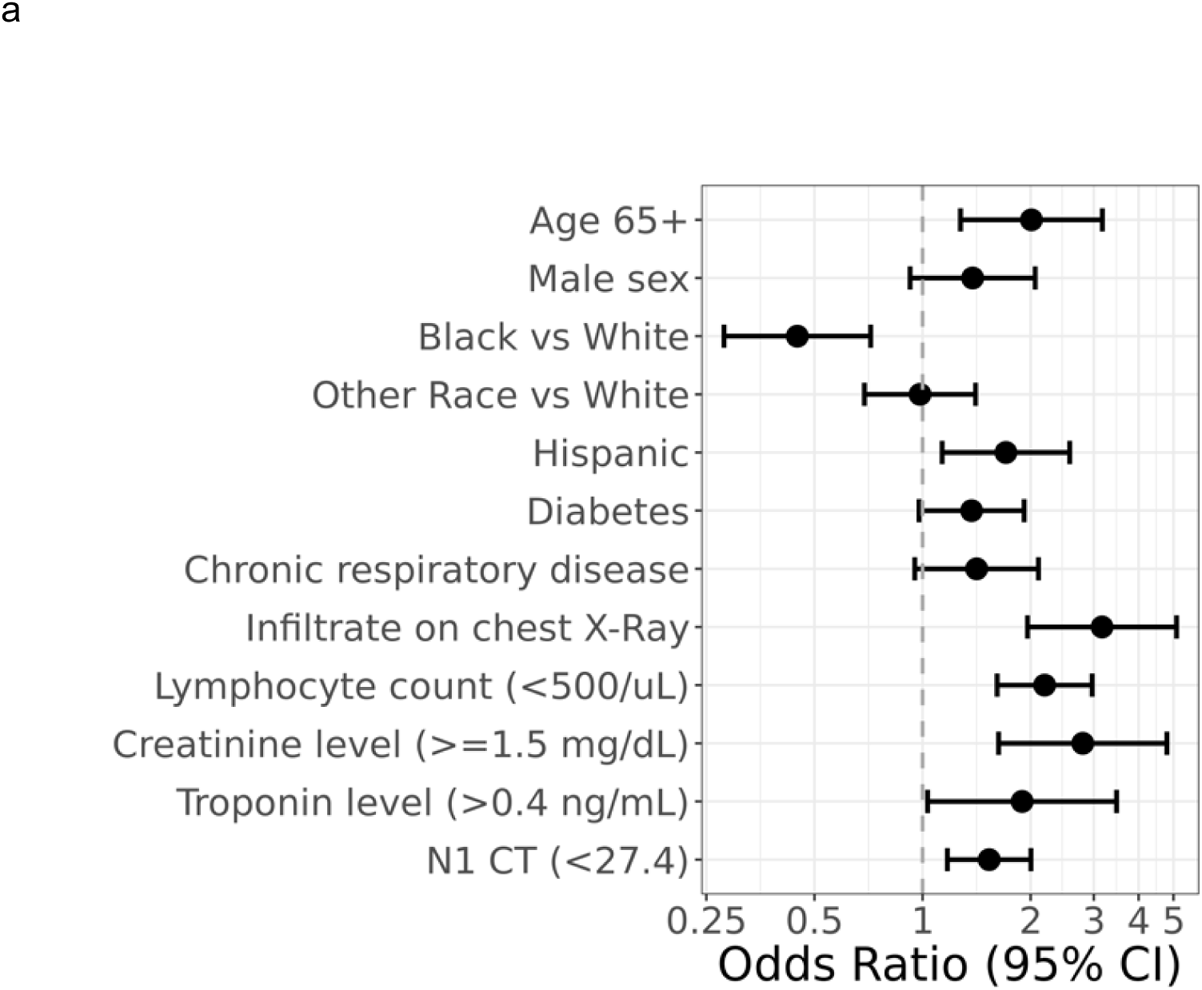
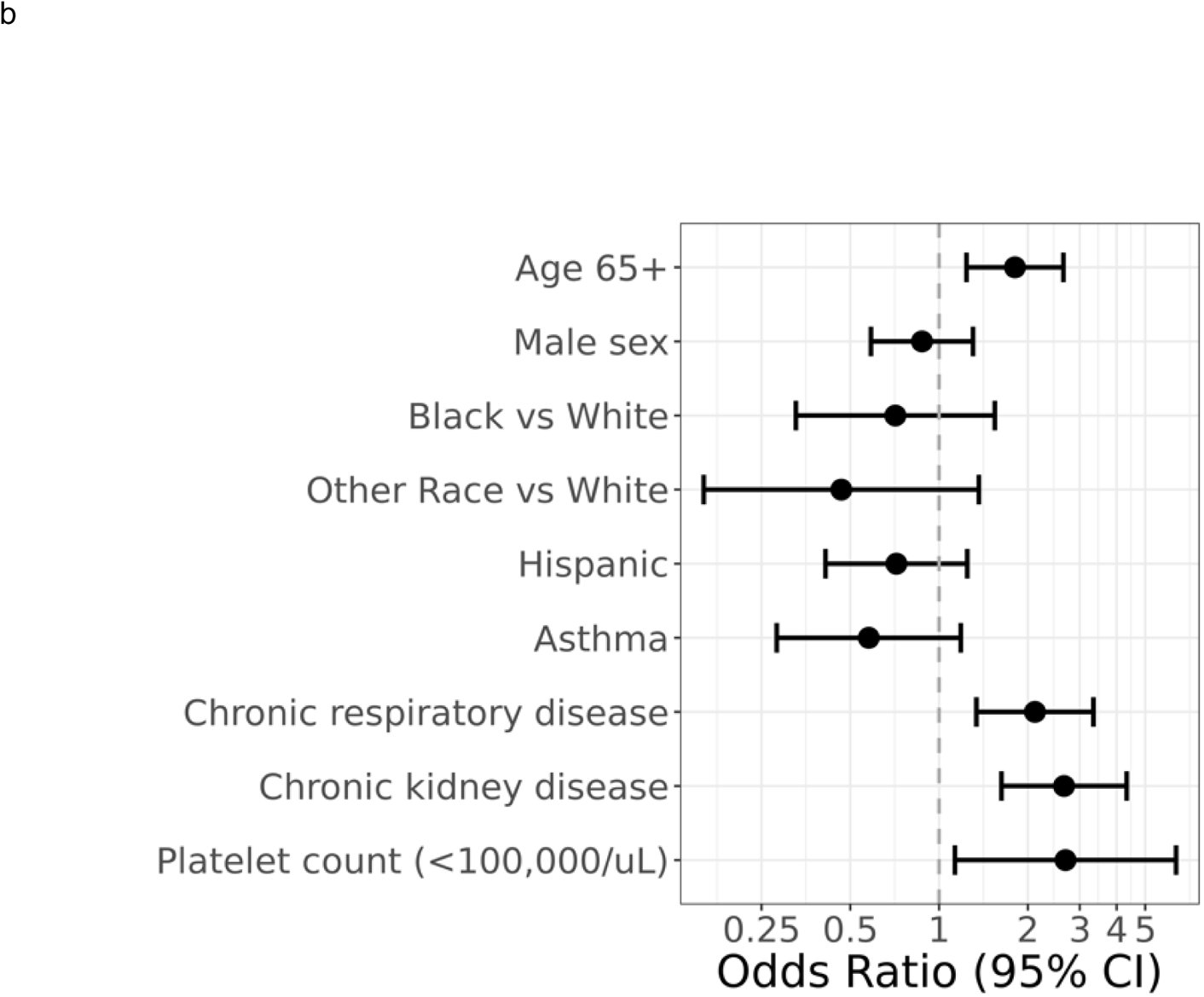
Forest Plot Models. (a) Adjusted odds ratios for factors associated with more severe (groups 4, 5) versus less severe (groups 1,2,3) disease. (N=1164) (b) Adjusted odds ratios for factors associated with 28-day mortality (group 5) versus prolonged hospitalized course (group 4) (N=320) Trajectory 1= brief length of stay; trajectory 2= intermediate length of stay; trajectory 3= intermediate length of stay with discharge limitations; trajectory 4= prolonged hospitalization; trajectory 5= fatal.

589 participants completed at least one quarterly survey post discharge and did not differ from non-respondents based on baseline demographics or period of enrollment but did per trajectory group assignment (TABLE 3 SUPPLEMENT). 108 deaths were reported prior to day 28 and 59 deaths were reported after day 28 mostly in patients in trajectory group 4 (n=36, 61%; p<0.001). Ten (2%) patients reported 11 SARS-CoV-2 reinfections post discharge. Vaccines were not available in the US for the general population until after study enrollment had largely been completed. 62% of patients reported receiving a COVID-19 primary vaccination series after discharge with 36% reporting booster doses, mostly with mRNA based vaccines (52% Pfizer BioNtech, 40% Moderna and 8% Johnson and Johnson for the primary series and 60% Pfizer BioNtech, 39% Moderna and 1% Johnson and Johnson products for the boost). 305 respondents (52%) reported symptoms after discharge. In those reporting any symptom after discharge, the median number of symptoms or organ systems affected was 1.0 (IQR=1.5). Among those reporting any symptoms, the symptoms were as follows: dyspnea (56%), muscle aches/myalgia (40%), cough (38%), headache (37%), fatigue/malaise (35%), loss of smell or taste (27%), red eye (26%), sore throat (14%), nausea or vomiting (13%) and fever, chills in less than 10%. The frequency of symptoms did not change during the follow up period (FIGURE 6 SUPPLEMENT). The presence of symptoms at follow up was only associated with female sex in a univariate analysis (p=0.02) and not with trajectory group, Ct values, or antibody levels (FIGURE 7 SUPPLEMENT).

## DISCUSSION

We identified five longitudinal clinical phenotypes based on respiratory OS in our cohort of hospitalized COVID-19 in hospitalized patients, and explored the relationship between demographic data, clinical features, readily available laboratory testing (immunologic, hematological, biochemical, and virologic), and radiographic findings with disease trajectories. Similar to other cohorts, we found that age ≥65 years,^8^ Latinx ethnicity, certain comorbidities, presence of chest radiographic infiltrate, and selected biomarkers at baseline were associated with more severe disease course, and worse outcomes. While many of these risk factors have previously been linked with mortality in other cohorts,^9–12^ our data is based on a multi-center prospective longitudinal study to comprehensively identify risk factors associated with trajectory of disease course over time. The clinical trajectory approach has advantages over the conventional cross-sectional approach by fully leveraging longitudinal data to analyze patient outcomes based on time in hospital, and disease severity.

In addition, our data demonstrate that higher SARS-CoV-2 viral burden at presentation is associated with worse disease severity, consistent with prior reports in small case series. ^12,13^ Our report is unique in validating this observation in a larger cohort, and in demonstrating the association between longitudinal assessment of viral load, and clinical disease course. Cohort patients with fatal outcome had delayed clearance of detectable virus by PCR as well as the lowest levels of anti-RBD, and anti-S IgG, suggesting that impaired functional antiviral immune response necessary to clear the virus, may play a key role in fatal cases.The described calculated ratio (IgG/Ct value) is unique compared to other studies,^15,16^ and may represent an accessible approach for patient risk stratification based on clinical laboratory testing.

The IMPACC cohort was diverse, with 31% participants being Hispanic/Latinx, and 22% black/African American, reflecting communities disproportionately affected by COVID-19^12^, and with wide geographic distribution within the USA. Consistent with prior reports, Hispanic/Latinx ethnicity was associated with increased risk for more severe disease, although ultimately neither race nor ethnicity were associated with death when assessing multivariable risk between the more severely ill groups (trajectories 4, and 5). Our data are consistent with prior retrospective cohorts which failed to demonstrate increased mortality in hospitalized disadvantaged minority populations^17^, unlike population based data on increased overall mortality in these communities.^12^

The overall 28-day mortality was 9%,and remained stable through the duration of the study. This finding is consistent with other reports^18^ where mortality remained relatively unchanged after the first 2 months of widespread circulation of SARS-CoV-2 in the US (March, and April 2020), but suggests no significant impact of care interventions for hospitalized patients with COVID-19 on mortality during the period of this study. In contrast to the hospitalized COVID-19 prospective patient cohort studied early during the pandemic in the UK (ISARIC WHO CCP-UK), our IMPACC study population showed lower overall mortality (9% vs 26%)^10^ though the characteristics of the study populations were different (with more major co-morbidities noted in the IMPACC cohort and an older age in the ISARIC WHO CCP-UK cohort).

Our prospective study did not demonstrate an association of obesity with poor outcome as was reported by others.^10,19^ This could in part be due to the fact that the majority of the patients (940; 81%) in IMPACC had a BMI above 25 kg/m^2^. It could be that impaired metabolic health^20^ (characterized by dyslipidemia, and insulin resistance) may be more predictive than BMI *per se*. A recent meta-analysis suggested that the effects of BMI on COVID-19 severity were less evident^21^. Additionally, although chronic respiratory disease tended to be more prevalent in patients with more severe illness, asthma was not a major confounding disease for COVID-19 severity in our cohort, possibly due to a previously reported protective role of type 2 immune inflammation^22^. Our ongoing longitudinal immunologic analyses may identify pathways distinguishing the impact of co-morbidities such as diabetes, obesity, and asthma on disease outcome from COVID-19.

While other small cohorts provide conflicting data on effects of antivirals^23^ and glucocorticoids^24^ on viral clearance, we did not find any relationship between viral clearance, and the use of remdesivir, or glucocorticoids^25^. Within the two most severe groups, viral RNA levels were highest in patients who received glucocorticoids, potentially reflecting nonrandom assignment of this therapy to more severe cases. In trajectory group 5, modest differences were observed in the slope of viral RNA levels potentially reflecting greater persistence of virus in individuals not treated with glucocorticoids, but this result is difficult to evaluate given that few patients were still alive at day 28.

Surveys conducted post-discharge in our study demonstrated that 51% of patients suffered from at least one symptom of PASC, which is in line with results from multiple other studies^26,27^. Female sex was a risk factor for PASC though the overall cohort was male predominant, reflecting a higher risk associated with male sex for hospitalization due to COVID-19. Though all symptoms improved compared to the acute presentation, conjunctivitis was more frequent at follow up than at presentation, as described by others ^28,29^. Some studies indicate that the risk of developing PASC may be diminished by vaccination^30^. However, our cohort completed enrollment prior to the nationwide rollout of vaccination, and therefore cannot address the characteristics of PASC in vaccine breakthrough cases^31^.

### Caveats and Limitations

Though IMPACC was racially, ethnically, and geographically diverse, the cohort excluded pregnant women known to be at risk for severe COVID-19^32^, and children with multisystem inflammatory syndrome.^33^ The cohort only recruited symptomatic, hospitalized patients and therefore our findings are not generalizable to asymptomatic or outpatient cases. We also did not collect information on socioeconomic status, thought to be a contributor of mortality in COVID-19.^34^ Additionally, PCR cannot distinguish between viable, and non-viable virus^35^. Prior studies have shown that the interval between symptom onset to viral clearance in culture did not exceed 12 days, and was much shorter than clearance of viral nucleic acid materials by PCR.^36^ In addition, particularly early in the pandemic, missed collection of nasal samples at days 14, and 28 in patients discharged from the hospital before these time points, and absence of further sampling once patients died could affect estimates of viral load, or serologies over time. While most participants were vaccinated by the time of follow-up; self-reporting of re-infection and small numbers precluded assessment of vaccine effectiveness. Our cohort was also fully enrolled prior to the widespread circulation of the SARS-CoV-2 B.1.617.2 (Delta), and B.1.1.529 (Omicron) variants which are associated with higher transmission, and possibly different severity of COVID-19. We assessed binding antibody levels rather than neutralizing antibody levels, though other studies have shown binding, and neutralizing antibody titers correlate well.^37^ Finally, the convalescent survey, planned at the beginning of the pandemic, did not include an extensive list of symptoms currently associated with PASC. While the severity of symptoms was not captured, ongoing efforts aim to analyze the impact of these persistent symptoms on quality of life, collected prospectively using patient-reported outcome measures.

### Conclusion

In this large, diverse, prospective cohort study of hospitalized patients in 20 sites across the U.S., we found that high baseline viral load, and its persistence were associated with more severe disease. Fatal cases were associated with the lowest levels of anti-RBD and S IgG. These findings suggest an impaired functional antiviral immune response necessary to clear the virus may play a key role in short-term mortality. The described calculated ratio (binding IgG/PCR Ct value) is unique compared to other studies, reflecting host pathogen interactions and appears to be an accessible approach for patient risk stratification. Immunophenotyping of blood, upper, and lower respiratory samples collected from this cohort is ongoing, and will enable identification of immune endotypes associated with severity of illness and/or persistence of symptoms. Furthermore, this effort will identify predictive and prognostic characteristics and generate hypotheses regarding the cellular and molecular basis of disease and recovery.

## CONTRIBUTORS

All authors read and approved the final version of the manuscript M.C.A, P.M.B and N.R. have verified the underlying data

− Conceptualization: A.O, P.M.B, M.C.A, N.R, A.A, L.B, C.S.C, G.M, C.B.C, V.S, C.L.H, K.C.N, J.S, A.C.S, D.C, N.I.A.H, C.B, S.B, E.M.
− Data curation: C.M, N.D.J, H.VB, S.L, F.KR
− Formal analysis: A.O, C.M, N.D.J, H.VB, M.C.A, D.E, S.L, F. KR
− Funding acquisition: O.L, R.S, E.H, A.F.S, W.M, M.D, B.PU, E.F, D.J.E, R.M, D.H, F.KH, J.M, M.K, M.A.A, L.I.E
− Investigation: L.B, C.S.C, G.M, C.B.C, N.R, V.S, C.H, K.C.N, J.M, A.C.S, D.C, N.I.A.H, C.B, S.B, E.M
− Methodology: A.O, C.M, N.D.J, H.VB, M.C.A, D.E, S.L, F.KR, P.M.B, N.R
− Project administration: P.M.B, A.A, A.O, S.K, B.PE, N.R, E.R, J.M
− Resources: A.O, J.S, N.D.J, C.M, C.S.C, C.B.C, M.K, L.B, A.C.S, F.KR, H.VB, D.E, S.L, A.F.S, V.S, D.H, R.M, S.K, O.L, C.B, E.H, D.E, B.PE, K.C.N, M.D, C.L.H, W.M, N.I.A.H, J.M, M.A.A, S.B, D.C, F.KH, L.I.E, E.M, G.M, R.S, J.D-A, B.PU, A.A, E.R, M.C.A, P.M.B, N.R
− Software: C.M, N.D.J, H.VB, S.L, A.O, J.M
− Supervision: P.M.B, A.O, M.C.A, N.R
− Validation: A.O, C.M, N.D.J, H.VB, M.C.A, D.E, S.L, F.KR
− Visualisation: A.O, C.M, N.D.J, H.VB
− Writing-original draft : A.O, C.M, M.C.A, J.S, N.R and P.M.B
− Writing review & editing: A.O, J.S, N.D.J, C.M, C.S.C, C.B.C, M.K, L.B, A.C.S, F.KR, H.VB, D.E, S.L, A.F.S, V.S, D.H, R.M, S.K, O.L, C.B, E.H, D.J.E, B.PU, R.K.N, M.D, C.H, W.M, N.I.A.H, J.M, M.A.A, S.B, D.C, F.KH, L.I.E, E.M, G.M, R.S, J.D-A, B.PE, A.A, E.R, M.C.A, P.M.B, N.R

Replication of analyses: A.O,C.M, N.D.J, M.C.A, H.VB

### IMPACC Study Group

*Steering Committee*: as above

*Clinical & Data Coordinating Center (CDCC):* Data curation

*IMPACC Data Analysis Group:* Formal analysis

*IMPACC Site Investigators:* Investigation

*IMPACC Core Labs:* Methodology

*IMPACC Study Team:* Investigation

## DECLARATION OF INTERESTS

J.S reports funds paid to institution by the National Institute of Health - National Institute of Allergy and Infectious Diseases for the completion of this manuscript.

C.B.C reports funds paid to institution from the National Institute of Health - National Institute of Allergy and Infectious Diseases for the completion of this manuscript. C.B.C also reports a grant paid to institution by the Bill & Melinda Gates Foundation for Covid-19 work. C.B.C is a consultant for bioMerieux on clinical biomarkers. He reports his participation on a data safety monitoring board or advisory board for the Convalescent plasma Covid-19 study for the National Heart, Lung and Blood Institute (NHLBI). CBC is also President, Board of Directors for the National Foundation of Emergency Medicine (NFEM), a non-profit supporting emergency medicine research and researchers.

M.K reports funds paid to institution for grants or contracts from the National Institute of Health.

L.B reports grant awarded to institution from the National Institute of Health - National Institute of Allergy and Infectious Diseases.

A.C.S reports funds paid to institution by the National Institute of Health for the completion of this manuscript (NIH U19 AI089992 – NIH K24 AG042489).

F.KR has received funds from the National Institute of Allergy and Infectious Diseases Collaborative Influenza Vaccine Innovation Centers (CIVIC) contract 75N93019C00051, the National Institute of Health - Centers of Excellence for influenza Research and Response, (CEIRR), 75N93021C00014, the JPB Foundation and the Open Philanthropy Project (research grant 2020-215611, 5384), the National Cancer Institute, National Institutes of Health, under Contract No. 75N91019D00024, Task Order No. 75N91020F00003 and Research funding from Pfizer for development of animal models for SARS-CoV-2. F.KR receives royalties from Avimex, and receives consulting fees from Pfizer, Seqirus, Avimex and Third Rock Ventures. F.KR has received several payments or honoraria for academic lectures over the past two years. F. KR is listed as a co-inventor on patents filed for applications relating to SARS-CoV-2 serological assays (the “Serology Assays”) and NDV-based SARS-CoV-2 vaccines.

H.vB reports receipt of the following grants paid to institution: National Institute of Health Centers of Excellence for influenza Research and Response, (CEIRR - 75N93021C00014), and National Institute of Health (Dengue Human Immunology Project Consortium - Mount Sinai IMPACC COVID-19 Cores - U19 AI118610 S1).

D. E reports funds paid to institution for grants or contracts from the National Institute of Health.

D.E is a statistician on the Data Safety Monitoring Board for the COLSTAT Trial. D.E is on the board of Directors for the Society for Clinical Trials.

V.S is listed as a co-inventor on a patent filed relating to SARS-CoV-2 serological assays (the “Serology Assays”).

R.M reports a grant from the National Institute of Health for the completion of this manuscript (AI 089992). R. M served as a Councilor for the Society of Leukocyte Biology from 2018 to 2021.

S.K is a personal consultant related to ImmPort data repository for Peraton.

O.L reports funds paid to institution by the National Institute of Health - National Institute of Allergy and Infectious Diseases for the completion of this manuscript (1-U19-AI118608-01A1).

O.L received payments in the past 36 months from Midsized Bank Coalition of America (MBCA) and Moody’s Analytics for presentations regarding the coronavirus pandemic.

D.J.E reports funds paid to institution for grants or contracts from the National Institute of Health.

K.C.N reports having received grants paid to institution from the following institutes in the past 36 months: National Institute of Allergy and Infectious Diseases (NIAID), National Heart, Lung, and Blood Institute (NHLBI), National Institute of Environmental Health Sciences (NIEHS), and Food Allergy Research & Education (FARE). K.C.N receives consulting fees from Excellergy, Eli Lilly, Red Tree Ventures, and Phylaxis. K.C.N has one Licensee: Alladapt and Before Brands (Application number: US15/048,609) for Mixed allergen composition and methods for using the same. K.C.N has two patents issued: Granulocyte-based methods for detecting and monitoring immune system disorders (Application number: US12/686,121), and Methods and Assays for Detecting and Quantifying Pure Subpopulations of White Blood Cells in Immune System Disorders (Application number: US12/610,94). K.C.N is the director of the World Allergy Organization Center of Excellence for Stanford and a co-founder of Seed Health, IgGenix, ClostraBio, and ImmuneID. K.C.N is also a co-founder of Before Brands, Alladapt, and Latitude and an advisor for Cour Pharma. K.C.N is a member of the national scientific committee for the Immune Tolerance Network (ITN) and the National Institutes of Health (NIH) clinical research centers.

C.L.H reports a grant awarded to institution by the National Institute of Health and the American Lung Association. C.L.H also reports having received consulting fees from the National Institute of Health. C.L.H has received payment for medical grand rounds at institution and support for attending meetings and/or travel from Critical care trialists, critical care reviews and multiple universities. C.L.H is on the data safety monitoring board of QuantumHealth for iSPY COVID and is on the board of directors for the American Thoracic Society.

W.M reports a grant from the National Institute of Health – National Institute of Allergy and Infectious Diseases for the completion of this manuscript (NIH NIAID R01AI14583).

J.P.M reports funds paid to institution for grants or contracts from the National Institute of Health for the completion of this manuscript (NIH Grant # 3U19AI062629-17S2).

M.A.A reports a grant from the National Institute of Health - National Institute of Allergy and Infectious Diseases for the completion of this manuscript (5U54AI142766-03).

L.E reports grant awarded to institution from the National Institute of Health (NIH R01AI104870-S1).

E.M reports funds paid to institution for grants or contracts from the National Institute of Health for the completion of this manuscript (NIH R01 AI104870S1). E.M also reports having received the following grants paid to institution in the past 36 months: the Babson Diagnostics Grant, the Austin Public Health Grant, and K08 26-1616-11. E.M received payments for lectures for the MS Association of America. E. M received payments for the participation in the advisory boards of Genentech, Horizon, Teva and Viela Bio.

G.M receives consulting fees from Gilead.

B.PE reports a grant from the National Institute of Health - National Institute of Allergy and Infectious Diseases for the completion of this manuscript.

E.F.R reports funds paid to institution by the National Institute of Health - National Institute of Allergy and Infectious Diseases for the completion of this manuscript (NIAID U19AI12891303).

M.C.A reports funds received from the National Institute of Health for the completion of this manuscript (NIH – R01AI32774) and funding by the National Institute of Allergy and Infectious Diseases for travel to present data related to this study.

P.M.B is a federal employee serving as project scientist for this project but has no role in funding decisions or oversight for relevant grants.

N.R reports funds paid to institution by the National Institute of Health - National Institute of Allergy and Infectious Diseases for the completion of this manuscript. N.R reports contracts with Lilly and Sanofi for conduct of Covid-19 clinical trials. N.R receives consulting fees from ICON EMMES for consulting on safety for Covid-19 clinical trials. N.R is an associate editor for Clinical infectious diseases.

The remaining authors declare that they have no conflict of interest.

## DATA SHARING STATEMENT

The IMPACC Data Sharing Plan is designed to enable the widest dissemination of data, while also protecting the privacy of the participants and the utility of the data by de-identifying and masking potentially sensitive data elements. This approach is fully compliant with the NIH public data sharing policy. The study protocol and clinical dataset are deposited at the Immunology Database and Analysis Portal (ImmPort), a NIAID Division of Allergy, Immunology, and Transplantation-funded data repository, under study accession SDY1760. After publication, it will be available to appropriate academic parties upon request and submission of a suitable study protocol, analysis plan, and signed data use agreement subject to NIAID approval via AccessClinicalData@NIAID (https://accessclinicaldata.niaid.nih.gov/study-viewer/clinical_trials). Please contact to view data for review purposes. All codes for the analyses and tables generated by this study are available in the <u>Bitbucket</u> repository.

## Data Availability

All data produced in the present study are available upon reasonable request to the authors
All data produced in the present work are contained in the manuscript

## IMPACC STUDY GROUP

### IMPACC Steering Committee

1. Clinical & Data Coordinating Center (CDCC); Precision Vaccines Program, Boston Children’s Hospital: Al Ozonoff, PhD, Joann Diray-Arce, PhD.
2. David Geffen School of Medicine at the University of California Los Angeles: Joanna Schaenman, MD, PhD, Elaine F. Reed, PhD.
3. Benaroya Research Institute: Matthew C. Altman, MD.
4. University of California San Francisco School of Medicine: Carolyn S. Calfee, MD, David J. Erle, MD.
5. Drexel University/Tower Health Hospital: Charles B. Cairns, MD, Elias K. Haddad, PhD.
6. University of Arizona: Monica Kraft, MD, Christian Bime, MD.
7. Boston Clinical Site: Precision Vaccines Program, Boston Children’s Hospital, Brigham and Women’s Hospital, and Harvard Medical School: Lindsey R. Baden, MD, Ofer Levy, MD, PhD.
8. Yale School of Medicine, and Yale School of Public Health: Albert C. Shaw, MD, PhD, David A. Hafler, MD, Ruth R. Montgomery, PhD, Steven H. Kleinstein, PhD.
9. Icahn School of Medicine at Mount Sinai: Ana Fernandez Sesma, PhD, Viviana Simon, MD, PhD.
10. Stanford University: Bali Pulendran, PhD, Kari C. Nadeau, MD, PhD, Mark M Davis, PhD.
11. Oregon Health Sciences University: Catherine L. Hough, MD, William B. Messer, MD, PhD.
12. Oklahoma University Health Sciences Center: Nelson I Agudelo Higuita, MD, Jordan P. Metcalf, MD.
13. University of Florida/University of South Florida: Mark A. Atkinson, PhD, Scott C. Brakenridge, MD.
14. Baylor College of Medicine, and the Center for Translational Research on Inflammatory Diseases, Michael E. DeBakey: David Corry, MD, Farrah Kheradmand, MD.
15. The University of Texas at Austin: Lauren I. R. Ehrlich, PhD, Esther Melamed, MD, PhD.
16. Case Western Reserve University: Grace A. McComsey, MD, Rafick Sekaly, PhD
17. La Jolla Institute for Immunology: Bjoern Peters, PhD.
18. National Institute of Allergy and Infectious Diseases/National Institutes of Health: Patrice M. Becker, MD, Alison D. Augustine, PhD.
19. Emory University: Nadine Rouphael, MD.

### Clinical & Data Coordinating Center (CDCC) (Precision Vaccines Program, Boston Children’s Hospital)

Al Ozonoff, PhD, Carly E. Milliren, MPH, Joann Diray-Arce, PhD, Kerry McEnaney, BS, Brenda Barton, BSN, RN, Claudia Lentucci, PhD, Mehmet Saluvan, PhD, Ana C. Chang, MS, Annmarie Hoch, BA, Marisa Albert, BSN, RN, Tanzia Shaheen, MS, MPH, Alvin T. Kho, PhD, Shanshan Liu, MS, MPH, Sanya Thomas, MBBS, Jing Chen, PhD, Maimouna D. Murphy, Mitchell Cooney, BA.

### IMPACC Data Analysis Group

1. Clinical & Data Coordinating Center (CDCC); Precision Vaccines Program, Boston Children’s Hospital: Al Ozonoff, PhD, Joann Diray-Arce, PhD.
2. David Geffen School of Medicine at the University of California Los Angeles.
3. Benaroya Research Institute: Naresh Doni Jayavelu, PhD, Matthew C. Altman, MD, Scott Presnell, PhD.
4. University of California San Francisco School of Medicine: Gabriela K. Fragiadakis, PhD, Ravi Patel, PhD.
5. Drexel University/Tower Health Hospital.
6. University of Arizona.
7. Precision Vaccines Program, Boston Children’s Hospital, Brigham and Women’s Hospital, and Harvard Medical School.
8. Yale School of Medicine, and Yale School of Public Health: Denise A. Esserman, PhD, Leying Guan, PhD, Steven H. Kleinstein, PhD, Jeremy Gygi, BS, Shrikant Pawar, PhD, Anderson Brito, PhD.
9. Icahn School of Medicine at Mount Sinai: Zain Khalil, MSc.
10. Stanford University.
11. Oregon Health Sciences University.
12. Oklahoma University Health Sciences Center.
13. University of Florida/University of South Florida.
14. Baylor College of Medicine, and the Center for Translational Research on Inflammatory Diseases, Michael E. DeBakey.
15. The University of Texas at Austin: Cole Maguire, BS.
16. Case Western Reserve University: Slim Fourati, PhD.
17. La Jolla Institute for Immunology: Bjoern Peters, PhD, James A. Overton, PhD, Randi Vita, MD, Kerstin Westendorf, PhD.
18. National Institute of Allergy and Infectious Diseases/National Institutes of Health.
19. Emory University.

### IMPACC Site Investigators

1. Clinical & Data Coordinating Center (CDCC): Precision Vaccines Program, Boston Children’s Hospital.
2. David Geffen School of Medicine at the University of California Los Angeles: Joanna Schaenman, MD, PhD, Ramin Salehi-Rad, MD, PhD.
3. Benaroya Research Institute.
4. University of California San Francisco School of Medicine: Carolyn S. Calfee, MD, David J. Erle, MD, Aleksandra Leligdowicz, MD, PhD, Michael A. Matthay, MD, Jonathan P. Singer MD, Kirsten N. Kangelaris, MD, Carolyn M. Hendrickson, MD, Matthew F. Krummel, PhD, Charles R. Langelier, MD, PhD, Prescott G. Woodruff, MD.
5. Drexel University/Tower Health Hospital: Charles B. Cairns, MD, Debra L. Powell, MD, James N. Kim, MD, Brent Simmons, MD, I. Michael Goonewardene, MD, Cecilia M. Smith, DO, Mark Martens, MD.
6. University of Arizona: Monica Kraft, MD, Christian Bime, MD, Jarrod Mosier, MD, Hiroki Kimura, MD, PhD.
7. Boston Clinical Site: Precision Vaccines Program, Boston Children’s Hospital, Brigham and Women’s Hospital, and Harvard Medical School: Lindsey R. Baden, MD, Amy C Sherman, MD, Stephen R Walsh, MD, Nicolas C Issa, MD.
8. Yale School of Medicine, and Yale School of Public Health: Albert C. Shaw, MD, PhD, Charles Dela Cruz, MD, PhD, Shelli Farhadian, MD, PhD, Akiko Iwasaki, PhD, Albert I. Ko, MD.
9. Icahn School of Medicine at Mount Sinai: Viviana Simon, MD, PhD.
10. Stanford University: Kari C. Nadeau, MD, PhD, Sharon Chinthrajah, MD, Neera Ahuja, MD, Angela J Rogers, MD, Maja Artandi, MD.
11. Oregon Health Sciences University: William B. Messer, MD, PhD, Sarah A.R. Siegel, PhD, MPH, Zhengchun Lu, MD, PhD.
12. Oklahoma University Health Sciences Center: Nelson I Agudelo Higuita, MD, Jordan P. Metcalf, MD, Douglas A. Drevets, MD, Brent R. Brown, MD.
13. University of Florida/University of South Florida: Scott C. Brakenridge, MD, Matthew L. Anderson, MD, PhD, Faheem W Guirgis, MD.
14. Baylor College of Medicine, and the Center for Translational Research on Inflammatory Diseases, Michael E. DeBakey: David Corry, MD, Farrah Kheradmand, MD.
15. The University of Texas at Austin: Esther Melamed, MD, PhD, Rama V Thyagarajan, MD, MPH, Justin F Rousseau, MD, MMSc, Dennis Wylie, PhD, Johanna Busch, MD, PhD, Saurin Gandhi, MD, Todd A. Triplett, PhD.
16. Case Western Reserve University: Grace A. McComsey, MD, George Yendewa, MD, Olivia Giddings, MD.
17. La Jolla Institute for Immunology.
18. National Institute of Allergy and Infectious Diseases/National Institutes of Health.
19. Emory University: Nadine Rouphael, MD, Evan J. Anderson, MD, Aneesh K. Mehta, MD, Jonathan E. Sevransky, MD.

### IMPACC Core Laboratory

1. Clinical & Data Coordinating Center (CDCC); Precision Vaccines Program, Boston Children’s Hospital: Joann Diray-Arce, PhD.
2. David Geffen School of Medicine at the University of California Los Angeles.
3. Benaroya Research Institute: Matthew C. Altman, MD, Bernard Khor, MD, PhD.
4. University of California San Francisco School of Medicine.
5. Drexel University/Tower Health Hospital.
6. University of Arizona.
7. Precision Vaccines Program, Boston Children’s Hospital, Brigham and Women’s Hospital, and Harvard Medical School.
8. Yale School of Medicine, and Yale School of Public Health: Ruth R. Montgomery, PhD.
9. Icahn School of Medicine at Mount Sinai: Florian Krammer, PhD, Harm van Bakel, PhD, Adeeb Rahman, PhD, Daniel Stadlbauer, PhD, Jayeeta Dutta, Hui Xie, MS, Seunghee Kim-Schulze, PhD, Ana Silvia Gonzalez-Reiche, PhD, Adriana van de Guchte, MS, Keith Farrugia, MSc, Zenab Khan, MSc.
10. Stanford University: Holden T. Maecker, PhD.
11. Oregon Health Sciences University.
12. Oklahoma University Health Sciences Center.
13. University of Florida/University of South Florida.
14. Baylor College of Medicine, and the Center for Translational Research on Inflammatory Diseases, Michael E. DeBakey.
15. The University of Texas at Austin.
16. Case Western Reserve University.
17. La Jolla Institute for Immunology.
18. National Institute of Allergy and Infectious Diseases/National Institutes of Health.
19. Emory University.

### IMPACC Study Team

1. Clinical & Data Coordinating Center (CDCC)
2. David Geffen School of Medicine at the University of California Los Angeles: David Elashoff, PhD, Jenny Brook, MS, Estefania Ramires-Sanchez, BS, Megan Llamas, BS, Adreanne Rivera, BS, Claudia Perdomo, AS, Dawn C. Ward, MD, Clara E. Magyar, PhD, Jennifer A. Fulcher, MD, PhD.
3. Benaroya Research Institute.
4. University of California San Francisco School of Medicine: Yumiko Abe-Jones, MS, Saurabh Asthana, PhD, Alexander Beagle, MD, Sharvari Bhide, BS, Sidney A. Carrillo, MPH, Suzanna Chak, BA, Gabriela K. Fragiadakis, PhD, Rajani Ghale, MPH, Ana Gonzalez, Alejandra Jauregui, Norman Jones, MS, Tasha Lea, MS, Deanna Lee, Raphael Lota, Jeff Milush, PhD, Viet Nguyen, BS, Logan Pierce, MD, Priya A. Prasad, PhD, Arjun Rao, PhD, Bushra Samad, MS, Cole Shaw, SM, Austin Sigman, BS, Pratik Sinha, MD, PhD, Alyssa Ward, PhD, Andrew Willmore, BS, Jenny Zhan, Sadeed Rashid, BS, Nicklaus Rodriguez, Kevin Tang, MS, Luz Torres Altamirano, BS, Legna Betancourt, Cindy Curiel, BA, Nicole Sutter, MPH, Maria Tercero Paz, BA, Gayelan Tietje-Ulrich, BA, Carolyn Leroux, BS, Ravi Patel, PhD.
5. Drexel University/Tower Health Hospital: Jennifer Connors, MSc, Mariana Bernui, PhD, Michel A. Kutzler, PhD, Carolyn Edwards, MSN, Edward Lee, BA, Edward Lin, BS, Brett Croen, BS, Nicholas C. Semenza, BS, Brandon Rogowski, BS, Nataliya Melnyk, BA, Kyra Woloszczuk, BA, Gina Cusimano, MSc, Mathew R. Bell, BS, Sara Furukawa, BA, Renee McLin, MSN, Pamela Marrero, CCRC, Julie Sheidy, BSN, George P. Tegos, PhD, Crystal Nagle, Nathan Mege, MSc, Kristen Ulring BS, MLS, Vicki Seyfert-Margolis, PhD.
6. University of Arizona: Michelle Conway, BS, Dave Francisco, MS, Allyson Molzahn, BS, Heidi Erickson, BSN, Connie Cathleen Wilson, MS, Ron Schunk, RT, Bianca Sierra, BS, Trina Hughes, AAS.
7. Boston Clinical Site: Precision Vaccines Program, Boston Children’s Hospital, Brigham and Women’s Hospital, and Harvard Medical School: Kinga Smolen, PhD, Michael Desjardins, MD, Simon van Haren, PhD, Xhoi Mitre, BS, Jessica Cauley, BS, Xiaofang Li, PhD, Alexandra Tong, BS, Bethany Evans, BS, Christina Montesano, BS, Jose Humberto Licona, MD, Jonathan Krauss, BS, Jun Bai Park Chang, BS, Natalie Izaguirre, BS.
8. Yale School of Medicine, and Yale School of Public Health: Omkar Chaudhary, PhD, Andreas Coppi, PhD, John Fournier, MD, Subhasis Mohanty, PhD, M. Catherine Muenker, MS, Allison Nelson, RN, Khadir Raddassi, PhD, Michael Rainone, William E. Ruff, PhD, Syim Salahuddin, MS, Wade L. Schulz, PhD, Pavithra Vijayakumar, MD, Haowei Wang, MD, PhD, Elsio Wunder Jr., DVM, PhD, H. Patrick Young, PhD, Yujiao Zhao, PhD.
9. Icahn School of Medicine at Mount Sinai: Miti Saksena, MBBS, Deena Altman, MD, Erna Kojic, MD, Komal Srivastava, MS, Lily Q Eaker, BA, Maria C Bermúdez-González, MPH, Katherine F Beach, BS, Levy A Sominsky, BA, Arman R. Azad, BA, Juan Manuel Carreño, PhD, Gagandeep Singh, PhD, Ariel Raskin, BA, Johnstone Tcheou, BS, Dominika Bielak, BA, Hisaaki Kawabata, BA Lubbertus CF Mulder, PhD, Giulio Kleiner, PhD.
10. Stanford University: Alexandra S. Lee, Evan Do, BS, Andrea Fernandes, MS, Monali Manohar, PhD, Thomas Hagan, PhD, Catherine A. Blish, MD, PhD, Hena Naz Din, MPH, Jonasel Roque, BS, Samuel Yang, MD.
11. Oregon and Health Sciences University: Amanda Brunton, MPH, Peter E. Sullivan, BA, Matthew Strnad, BA, Zoe L. Lyski, MS, Felicity J. Coulter, MSc.
12. Oklahoma University Health Sciences Center: J. Leland Booth, MS, Lauren A. Sinko, RN, BSN.
13. University of Florida/University of South Florida: Lyle L. Moldawer, PhD, Brittany Borresen, MSc, Brittney Roth-Manning, MPH.
14. Baylor College of Medicine, and the Center for Translational Research on Inflammatory Diseases, Michael E. DeBakey: Li-Zhen Song, MD, Ebony Nelson, MS.
15. The University of Texas at Austin: Megan Lewis-Smith, MD, Jacob Smith, MD, Cole Maguire, BS, Pablo Guaman Tipan, BS, Nadia Siles, BS, Sam Bazzi, BS, Janelle Geltman, MS, Kerin Hurley, RN, Gio Gabriele, BS.
16. Case Western Reserve University: Scott Sieg, PhD.
17. La Jolla Institute for Immunology:
18. National Institute of Allergy and Infectious Diseases/National Institutes of Health: Tatyana Vaysman, MD.
19. Emory University: Laurel Bristow, MSc, Laila Hussaini, MPH, Kieffer Hellmeister, BS, Hady Samaha, MD, Andrew Cheng, BA, Christine Spainhour, RN, Erin M. Scherer, PhD, DPhil, Brandi Johnson, BS, Amer Bechnak, MD, Caroline R. Ciric, BS, Lauren Hewitt, Erin Carter, MS, Nina Mcnair, BS, Bernadine Panganiban, BS, Christopher Huerta, BS, Jacob Usher, BS, Susan Pereira Ribeiro, PhD.

## ACKNOWLEDGMENTS

Shun Rao, BS and Sofia Vignolo, BS, Brandon Larson, Jane Buckner, Hiromitsu Asashima, Natasha Balkcom, Santos Bermejo, Ryan Borg, Kristina Brower, Arnau Casanovas-Massana, Michael Chiorazzi, Erendira Di Giuseppe, Coriann Dorgay, Dimitri Duvilaire, Rebecca Earnest, Brinda Emu, Nathan Grubaugh, Roy S. Herbst, Maxine Kuang, Sarah Lapidus, Zitong Lin, Yicong Liu, Maksym Minasyan, Adam J. Moore, Alexander Robertson, Denise Shepard, Xiaomei Wang, Elizabeth B. White, Anne L. Wyllie, Emily D. Ferreri, Rachel L. Chernet, Ashley-Beathrese Salimbangon, Charles Gleason, Steve Sigelman, Julia Goldstein, Ari Joffe, Alkis Togias, Daniel Rotrosen, Tigisty Girmay, Lisa Harewood, Cecilia Zhang, Jessica Traenkner, Cindy Lubbers, Meg Taylor, Natalie Gray, Dongli Wang, Juliet Morales, Mai Biak Kio, Youssef Saklawi, David Constant, Clive Woffendin, Akram Khan, Maria Cristina Crizaldo, Jennifer Lanz, Richard MadolidKieran McGrail, Sean McGrail, Melissa Terry-White, Ricardo Ungaro, MacKenzie Williams, Elaine Ramirez, Jacob Rogers, Yousuf Ahmed, Coburn Allen, Michael Brode, Apoorva Kakkilaya, Nisha Holay, Blaine Caslin, Sneha Banerjee, Maisey Schuler, John Moore, Parker Davis, Mary Kelley.

Arash Naeim, Marianne Bernardo, Sarahmay Sanchez, Shannon Intluxay, Azlann Arnett, Kimberly Yee, Douglas Zhang, Maria Calvo, Jeremy Giberson, Serena Ke, Cathy Cai, Pat Glenn, Kristine Wong, Andra Blomkalns, Ruth O’Hara, Mike Leipold, Yael Rosenberg-Hasson, Corinne Fargo, Jessica Smith, David X. Lee, Olivia Krol, Lili Tian, Wenxin Wu, Kassidy Malone, Hayden Younger, Rachel Karlnoski, Naya Martin, Jordan Oberhaus, Maja Okuka, Travis Roundtree, Suzane Silbert, Thanh Tran, Mary Consolo, and Heather Tribout.

## Funding

The study was funded by the United States National Institutes of Health through the following grants: 5R01AI135803-03, 5U19AI118608-04, 5U19 AI128910-04, 4U19AI090023-11, 4U19AI118610-06, R01AI145835-01A1S1, 5U19AI062629-17, 5U19AI057229-17, 5U19AI125357-05, 5U19AI128913-03, 3U19AI077439-13, 5U54AI142766-03, 5R01AI104870-07, 3U19AI089992-09, 3U19AI128913-03

## Notes

### Funding Statement

The study was funded by the United States National Institutes of Heath through the following grants: 5R01AI135803-03, 5U19AI118608-04, 5U19 AI128910-04, 4U19AI090023-11, 4U19AI118610-06, R01AI145835-01A1S1, 5U19AI062629-17, 5U19AI057229-17, 5U19AI125357-05, 5U19AI128913-03, 3U19AI077439-13, 5U54AI142766-03, 5R01AI104870-07, 3U19AI089992-09

### Author Declarations

IRB of University of Texas at Austin gave ethical approval of this work: IRB 2020-04-0117 IRB of University of California San Francisco gave ethical approval for this work: IRB 20-30497 IRB of Case Western reserve university gave ethical approval for this work: IRB STUDY20200573 The Department of Health and Human Services Office for Human Research Protections (OHRP) concurred that the study satisfied criteria for the public health surveillance exception and twelve institutions elected to conduct the study as public health surveillance.

