## Supplementary material for "Phenotypes of disease severity in a cohort of hospitalized COVID-19 patients: results from the IMPACC study": Online supplemental material

ONLINE SUPPLEMENT

*Study Participants*

Several site-dependent methods were used to identify eligible participants, including (but not limited to) acquisition of hospital-based service lists, and notification of research team of any positive SARS-CoV-2 PCR testing.

*Data Collection, Study Variables, and Biologic Samples*

Additional samples were collected within 24, and 96 hours of any care escalation to ICU-level care, or readmission after hospital discharge.

Escalation of care was defined as below:

 1. Transfer to an ICU, or ICU‐level of care

2. Initiation of any of the following interventions:

Respiratory failure requiring the initiation of invasive mechanical ventilation, or ECMO

Shock defined as hypotension requiring vasopressor support (> 5 mcg/kg/minute of dopamine, or equivalent)

3. Study patient was discharged, and then re‐admitted > 24 hours after discharge.

The following symptoms were surveyed at 3, 6, 9 and 12 months after discharge:

- Upper respiratory symptoms: sore throat, conjunctivitis/red eyes
- Cardiopulmonary symptoms: shortness of breath (dyspnea), cough
- Systemic symptoms: fever, chills, fatigue/malaise, muscle aches (myalgia)
- Neurologic symptoms: loss of smell/taste (anosmia), headache
- Gastrointestinal symptoms: nausea/vomiting

At each quarterly symptom survey, patients were also asked if they had been diagnosed with a new COVID-19 infection (Y/N), and if they had received a COVID vaccination since their last visit (if so which vaccine). Three COVID-19 vaccines were authorized/approved in the US by April 2022:

- Pfizer/BioNTech BNT162b2 mRNA vaccine
- ModernaTX, Inc mRNA‑1273 vaccine
- Janssen Pharmaceutical/Johnson & Johnson adenovirus Ad26.COV.2 vaccine

Survey was collected via app or telephone or in person visits by study coordinator. A secure electronic data collection form was designed to collect all relevant deidentified variables. Processed samples were barcoded, and centrally tracked on a laboratory information management system (LDMS, Frontier Science). All data were reviewed centrally to ensure accuracy, and consistency. Any data concerns were resolved by querying the site.

*SARS-CoV-2 PCR*

To assess SARS-CoV-2 viral load, nasal swabs were collected, and placed in 1ml of Zymo-DNA/RNA shield reagent (Zymo Research). RNA was extracted using the quick DNA-RNA MagBead kit (Zymo Research) following the manufacturer’s instructions. RT-PCR for SARS-CoV-2 was performed on RNA extracts using the SARS-CoV-2 (2019-nCoV) CDC qPCR Probe Assay with target genes 2019-nCoV_N1, 2019-nCoV_N2, and control human RNase P (Integrated DNA Technologies)^10^. A ceiling cutoff of 40 cycles was applied as the limit of detection. Samples with an undetectable human RNase P were considered collection failures, and were excluded.

*SARS-CoV-2 Sequencing*

*RNA-sequencing cDNA Library Production*

From each nasal RNA sample, 10µl was aliquoted to a library construction plate using the Perkin Elmer Janus Workstation (Perkin Elmer, Janus II).  Ribosomal depletion, cDNA synthesis and library construction steps were performed using the Total Stranded RNA Prep with Ribo-Zero Plus kit following the manufacturer’s instructions (Illumina). All steps were automated on the Perkin Elmer Sciclone NGSx Workstation to reduce batch to batch variability and to increase sample throughput. Final cDNA libraries were quantified using the Quant-it dsDNA High Sensitivity assay, and library insert size distribution was checked using a fragment analyzer (Advanced Analytical; kit ID DNF474). Samples where adapter dimers constituted more than 4% of the electropherogram area were failed prior to sequencing. Technical controls (K562,Thermo Fisher Scientific, cat# AM7832) were compared to expected results to ensure that batch to batch variability was minimized. Successful libraries were normalized to 10nM for sequencing.

*RNA-sequencing Clustering and Sequencing*

Barcoded libraries were pooled using liquid handling robotics prior to loading. Massively parallel sequencing-by-synthesis with fluorescently labeled, reversibly terminating nucleotides was carried out on the NovaSeq 6000 sequencer using S4 flowcells with target depth of 50 million 100 base-pair paired-end reads per sample (25 million read pairs).

*Nasal viral genome sequencing and assembly*

For viral genome sequencing, cDNA synthesis was performed with random hexamers and ProtoScript II (New England Biolabs, E6560) starting from 7 ul of total RNA extracted from clinical specimens. The SARS-CoV-2 genome was then amplified with Q5 Hot Start High-Fidelity DNA polymerase (New England Biolabs, cat. M0493) using two sets of custom designed tiling primers generating overlapping amplicons of ~1.5 and 2 Kb. The PCR amplification parameters were: 1 min at 98C, 35 cycles of 15s at 98C and 5 min at 63C, and final extension for 10 min at 65C. After equivolume pooling of the amplicons and cleanup with 1.8X volume of Ampure XT beads, libraries were prepared using the Nextera XT DNA Sample Preparation kit (Illumina, FC-131-1096), followed by paired-end sequencing (2x150nt) on the Illumina MiSeq platform. A custom reference-based analysis pipeline, https://github.com/mjsull/COVID_pipe, was used to assemble SARS-CoV-2 genomes.

*Serology*

Two readouts were generated for serology measures, endpoint titers, and area under the curve (AUC) values. These two readouts generally correlate well. However, AUC take not just the endpoint titer of ELISA curves but also the magnitude of the signal at each dilution into account, and therefore provides more granularity. For this reason, AUC was chosen as the main readout.

*Statistical analysis*

*Determination of Trajectory Groupings*

Ordinal scores corresponding to death (OS=7) were extrapolated to all subsequent scheduled visits after the date of death through day-28 for the purposes of modeling; ordinal scores corresponding to discharge (OS=1,2) were extrapolated to subsequent scheduled visits through day-28, or until a readmission was recorded. Any missing ordinal scores for remaining inpatient visits were filled in using a last visit carried forward approach with the exception of participant withdrawal where information was only carried forward until the date of withdrawal. For each specification of group number, polynomial curves up to cubic degree were allowed for each group, and polynomial order for each cluster was determined by backwards selection using retention threshold of p<0**^.^**05. All models included timing of symptom onset relative to hospital admission as fixed effect. The best-fitting model with respect to number of clusters, and polynomial order was selected using Akaike Information Criteria (AIC).

*Logistic Regression Modeling*

Logistic mixed-effect models were used to associate clinical characteristics, and laboratory values with disease severity expressed as different dichotomizations of the five-level clinical trajectory groups. All models included random intercept by enrollment site. A first set of models identified factors associated with less severe (trajectories 1, 2, and 3) versus more severe disease (trajectories 4, and 5) using the full cohort, while the second set of models identified factors associated with 28-day mortality (trajectory 5) within the subset with severe disease. Blocks of predictor variables were entered into the model sequentially: demographic factors, followed by clinical comorbidities at admission, other clinical factors including respiratory support, and radiographic findings at baseline visit, laboratory values at baseline visit, baseline viral load, and anti-RBD IgG dichotomized at the median. Clinical comorbidities considered for inclusion in the models were those with p-value<0**^.^**10 across groups in bivariate analysis, and overall prevalence of 10%, or greater. Backward selection was used to select a final model, starting with the full set of model predictors, and removing predictors with the highest p-value first until all p<0**^.^**20 with the exception of sociodemographic factors, which were forced into the model regardless of significance. Logistic mixed-effects models were fit with random intercept by clinical recruitment site. Beta coefficients are expressed as odds ratios (ORs) with 95% confidence intervals.

*Ordinal/Logistic Regression Modeling for Viral Load*

We performed ordinal, and logistic regression with mixed-effects using clmm, and lmer to model the association between baseline viral load, and clinical trajectories, after accounting for random enrollment site effect, and fixed age, and sex effects.  For the ordinal regression model, we treat the clinical trajectory from 1 to 5 as ordered, and representing an increase in the disease severity. Two-group comparisons are also carried out using logistic regression where we compare each pair of clinical trajectories to identify non-global patterns.

*Generalized Additive Regression Modeling for Viral Load*

We applied mixed-effect generalized additive model using the “mgcv” package in R to model SARS-CoV2 Ct values, and anti-viral IgG serology values for each trajectory group for samples collected within 28 days of hospital admission. Each model is adjusted for fixed effects of age, and sex, and random effects for participant, and enrollment site. Where indicated, models were adjusted for administration of remdesivir, or glucocorticoid at any time during hospitalization, and where indicated, trajectory groups were subdivided based on administration of one of these medications.

Figure legends

FIGURE 1 SUPPLEMENT

(a) Kaplan Meier survival curve

(b) Mortality per quarter

(c) Length of stay (with death as competing risk)

(d) Length of hospital stay (censored for death) by trajectory group [Trajectory 1= brief length of stay (green); trajectory 2= intermediate length of stay (teal); trajectory 3= intermediate length of stay with discharge limitations (blue); trajectory 4= prolonged hospitalization (orange); trajectory 5= fatal (red)].

FIGURE 2 SUPPLEMENT

(a) N2 Ct baseline: Shown are the N2 Ct values during the first study collection timepoint which occurred at any time from admission until up to approximately 48 hours after admission. Shown are median values (horizontal lines), mean values (black points), interquartile ranges (boxes), and 1**^.^**5 IQR (whiskers), as well as all individual points.

(b) N2 Ct over time: Shown are the N2 Ct values collected during the first 28 days of hospital admission. Trend lines for each trajectory represent the fit of a generalized additive model (GAM) to capture non-linear trends in the data.

(c) Anti-spike IgG titers at baseline: Shown are the anti-spike IgG values during the first study collection timepoint which occurred at any time from admission until up to approximately 48 hours after admission. Shown are median values (horizontal lines), mean values (black points), interquartile ranges (boxes), and 1**^.^**5 IQR (whiskers), as well as all individual points.

(d) Anti-spike IgG over time: Shown are the anti-spike IgG values collected during the first 28 days of hospital admission. Trend lines for each trajectory represent the fit of a generalized additive model (GAM) to capture non-linear trends in the data.

(e) Ratio anti-spike IgG/N1 Ct over time: Shown are the trends of the scaled ratio anti-Spike IgG values divided by scaled SARS-CoV-2 N1 gene Ct values for time points collected during the first 28 days of hospital admission, emphasizing the divergence of trajectory 5 from the other 5 trajectories in this measure.

Trajectory 1= brief length of stay (green); trajectory 2= intermediate length of stay (teal); trajectory 3= intermediate length of stay with discharge limitations (blue); trajectory 4= prolonged hospitalization (orange); trajectory 5= fatal (red).

FIGURE 3 SUPPLEMENT

(a) N1 Ct over time by remdesivir treatment: Shown are the SARS-CoV-2 N1 gene Ct values collected during the first 28 days of hospital admission. Each trajectory group is split into individuals who were, or were not treated with remdesivir during hospitalization. Trend lines for each group are represent the fit of a generalized additive model (GAM) to capture non-linear trends in the data. Shown are the p-values for the intercept, and slope comparing within each trajectory group those individuals who were (blue) vs were not (gray) treated with remdesivir.

(b) N1 Ct over time by glucocorticoid treatment: Shown are the SARS-CoV-2 N1 gene Ct values collected during the first 28 days of hospital admission. Each trajectory group is split into individuals who were, or were not treated with glucocorticoids during hospitalization. Trend lines for each group are represent the fit of a generalized additive model (GAM) to capture non-linear trends in the data. Shown are the p-values for the intercept, and slope comparing within each trajectory group those individuals who were (pink) vs were not (gray) treated with glucocorticoids.

FIGURE 4 SUPPLEMENT

(a) N1 Ct by Symptom Onset Days: Shown are the SARS-CoV-2 N1 gene Ct values per symptoms onset days. Trend lines for each trajectory represent the fit of a generalized additive model (GAM) to capture non-linear trends in the data.
(b) N2 Ct by Symptom Onset Days: Shown are the SARS-CoV-2 N2 gene Ct values per symptoms onset days. Trend lines for each trajectory represent the fit of a generalized additive model (GAM) to capture non-linear trends in the data.
(c) Anti-RBD IgG Titers by Symptom Onset Days: Shown are the anti-RBD IgG values per symptoms onset days (0-7 days; 7-14 days, and more than 14 days). Shown are median values (horizontal lines), mean values (black points), interquartile ranges (boxes), and 1**^.^**5 IQR (whiskers), as well as all individual points.

(d) Anti-spike IgG Titers by Symptom Onset Days: Shown are the anti-S IgG values per symptoms onset days (0-7 days; 7-14 days, and more than 14 days). Shown are median values (horizontal lines), mean values (black points), interquartile ranges (boxes), and 1**^.^**5 IQR (whiskers), as well as all individual points.

Trajectory 1= brief length of stay (green); trajectory 2= intermediate length of stay (teal); trajectory 3= intermediate length of stay with discharge limitations (blue); trajectory 4= prolonged hospitalization (orange); trajectory 5= fatal (red).

FIGURE 5 SUPPLEMENT

Identification of Pango lineages viral sequencing by

(a) Month of enrollment

(b) Clinical trajectory

Trajectory 1= brief length of stay (green); trajectory 2= intermediate length of stay (teal); trajectory 3= intermediate length of stay with discharge limitations (blue); trajectory 4= prolonged hospitalization (orange); trajectory 5= fatal (red).

FIGURE 6 SUPPLEMENT

Frequency of symptoms reported at baseline and 3,6,9 and 12 months after discharge in survey respondents.

FIGURE 7 SUPPLEMENT

(a) Shown are the SARS-CoV-2 N1 gene Ct values during the first study collection timepoint which occurred at any time from admission until up to approximately 48 hours after admission. Shown are median values (horizontal lines), mean values (black points), interquartile ranges (boxes), and 1**^.^**5 IQR (whiskers), as well as if symptoms were present (blue) or absent (red) at follow up.

(b) N1 Ct over 28 days and presence or absence of symptoms during follow up: Shown are the SARS-CoV-2 N1 gene Ct values collected during the first 28 days of hospital admission. Participants are split into individuals who eventually had persistent or new symptom (blue) or not (red) at follow up. Trend lines for each group are represent the fit of a generalized additive model (GAM) to capture non-linear trends in the data. Shown are the p-values for the intercept, and slope comparing those individuals who did (blue) vs did not (red) have symptoms at follow up.

(c) Shown are the RBD IgG during the first study collection timepoint which occurred at any time from admission until up to approximately 48 hours after admission. Shown are median values (horizontal lines), mean values (black points), interquartile ranges (boxes), and 1**^.^**5 IQR (whiskers), as well as if symptoms were present (blue) or absent (red) at follow up.

(d) RBD Ig G over 28 days and presence or absence of symptoms during follow up: Shown are the RBD IgG values collected during the first 28 days of hospital admission. Participants are split into individuals who eventually had persistent or new symptom (blue) or not (red). Trend lines for each group are represent the fit of a generalized additive model (GAM) to capture non-linear trends in the data. Shown are the p-values for the intercept, and slope comparing those individuals who did (blue) vs did not (red) have symptoms at follow up.

TABLE 1 SUPPLEMENT

Complications during hospitalization (N=1,164)

|  | Overall  (n=1,164) | Trajectory 1  (n=258) | Trajectory 2  (n=310) | Trajectory 3  (n=276) | Trajectory 4  (n=212) | Trajectory 5  (n=108) | Overall  p-value |
| --- | --- | --- | --- | --- | --- | --- | --- |
| Number of complications, median (IQR) | 2 (2) | 1 (2) | 2 (2) | 2 (2) | 3**^.^**5 (4) | 5 (4) | <**^.^**001 |
| Any complications, No. (%) | 958 (82) | 184 (71) | 239 (77) | 234 (85) | 197 (93) | 104 (96) | <**^.^**001 |
| Acute renal injury/ failure | 249 (21) | 28 (11) | 44 (14) | 46 (17) | 70 (33) | 61 (56) | <**^.^**001 |
| Shock (use of vasopressors) | 173 (15) | 3 (1) | 10 (3) | 10 (4) | 80 (38) | 70 (65) | <**^.^**001 |
| Anemia | 161 (14) | 15 (6) | 17 (5) | 32 (12) | 69 (33) | 28 (26) | <**^.^**001 |
| Liver dysfunction/ failure | 122 (10) | 18 (7) | 28 (9) | 33 (12) | 31 (15) | 12 (11) | 0**^.^**071 |
| Bacteremia | 113 (10) | 5 (2) | 16 (5) | 19 (7) | 45 (21) | 28 (26) | <**^.^**001 |
| Atrial fibrillation | 87 (7) | 5 (2) | 17 (5) | 19 (7) | 22 (10) | 24 (22) | <**^.^**001 |
| Congestive heart failure, or cardiomyopathy | 77 (7) | 7 (3) | 15 (5) | 19 (7) | 12 (6) | 24 (22) | <**^.^**001 |
| Venous thromboembolism | 75 (6) | 2 (1) | 10 (3) | 11 (4) | 36 (17) | 16 (15) | <**^.^**001 |
| Bleeding | 65 (6) | 5 (2) | 7 (2) | 9 (3) | 23 (11) | 21 (19) | <**^.^**001 |
| Hyperglycemia | 41 (4) | 4 (2) | 10 (3) | 15 (5) | 8 (4) | 4 (4) |  |
| Ventricular arrhythmia | 36 (3) | 1 (0) | 7 (2) | 2 (1) | 13 (6) | 13 (12) | <**^.^**001 |
| Pneumothorax | 32 (3) | 1 (0) | 4 (1) | 5 (2) | 15 (7) | 7 (6) | <**^.^**001 |
| Non-ST elevation myocardial infarction, or other cardiac ischemia | 28 (2) | 7 (3) | 1 (0) | 5 (2) | 5 (2) | 10 (9) | <**^.^**001 |
| Coagulation disorder / disseminated intravascular coagulation | 22 (2) | 3 (1) | 6 (2) | 1 (0) | 7 (3) | 5 (5) | 0**^.^**028 |
| Other complications | 20 (2) | 3 (1) | 5 (2) | 5 (2) | 5 (2) | 2 (2) | 0**^.^**905 |
| Stroke/ cerebrovascular accident | 19 (2) | 1 (0) | 0 (0) | 3 (1) | 10 (5) | 5 (5) | <**^.^**001 |
| ST-elevation myocardial infarction | 12 (1) | 3 (1) | 0 (0) | 1 (0) | 4 (2) | 4 (4) | 0**^.^**009 |
| Pneumomediastinum | 12 (1) | 0 (0) | 0 (0) | 0 (0) | 7 (3) | 5 (5) | <**^.^**001 |
| Meningitis | 11 (1) | 1 (0) | 1 (0) | 1 (0) | 4 (2) | 4 (4) | 0**^.^**008 |
| Arterial thromboembolism excluding stroke/CVA, or myocardial infarction | 10 (1) | 1 (0) | 1 (0) | 1 (0) | 5 (2) | 2 (2) | 0**^.^**053 |
| Bronchiolitis | 6 (1) | 0 (0) | 1 (0) | 1 (0) | 3 (1) | 1 (1) | 0**^.^**25 |
| Pancreatitis | 6 (1) | 4 (2) | 0 (0) | 1 (0) | 1 (0) | 0 (0) | 0**^.^**104 |
| Myocarditis/pericarditis | 5 (0) | 1 (0) | 2 (1) | 2 (1) | 0 (0) | 0 (0) | 0**^.^**683 |
| Seizures | 5 (0) | 1 (0) | 1 (0) | 1 (0) | 1 (0) | 1 (1) | 0**^.^**945 |
| Endocarditis | 1 (0) | 0 (0) | 0 (0) | 0 (0) | 1 (0) | 0 (0) | 0**^.^**343 |

p-value from Kruskal-Wallis test for number of complications, and chi-square test for all others.

Trajectory 1= brief length of stay; trajectory 2= intermediate length of stay; trajectory 3= intermediate length of stay with discharge limitations; trajectory 4= prolonged hospitalization; trajectory 5= fatal.

TABLE 2 SUPPLEMENT

Medication use during hospitalization (N=1,164)

| Drug, No. (%) | Overall  (n=1,164) | Trajectory 1  (n=258) | Trajectory 2  (n=310) | Trajectory 3  (n=276) | Trajectory 4  (n=212) | Trajectory 5  (n=108) | Overall  p-value |
| --- | --- | --- | --- | --- | --- | --- | --- |
| Remdesivir | 725 (62) | 115 (45) | 220 (71) | 171 (62) | 155 (73) | 64 (59) | <**^.^**001 |
| Steroids | 791 (68) | 127 (49) | 198 (64) | 192 (70) | 186 (88) | 88 (81) | <**^.^**001 |
| Other immunomodulators^1^ | 37 (3) | 2 (1) | 8 (3) | 4 (1) | 13 (6) | 10 (9) | <**^.^**001 |
| Azithromycin | 354 (30) | 57 (22) | 91 (29) | 77 (28) | 91 (43) | 38 (35) | <**^.^**001 |
| Other Antibiotics | 643 (55) | 109 (42) | 143 (46) | 129 (47) | 168 (79) | 94 (87) | <**^.^**001 |
| Convalescent Plasma | 99 (9) | 8 (3) | 20 (6) | 24 (9) | 31 (15) | 16 (15) | <**^.^**001 |
| Monoclonal Antibodies | 6 (1) | 0 (0) | 2 (1) | 1 (0) | 3 (1) | 0 (0) | 0**^.^**242 |
| Immunosuppressives | 80 (7) | 16 (6) | 17 (5) | 22(8) | 19 (9) | 6(6) | 0^.^497 |
| Hydroxychloroquine | 19 (2) | 5 (2) | 6 (2) | 3 (1) | 3 (1) | (2) | 0**^.^**92 |

^1^Anti-IL6, Anti-CCR5, Jak Inhibitors

p-value from chi-square test.

Trajectory 1= brief length of stay; trajectory 2= intermediate length of stay; trajectory 3= intermediate length of stay with discharge limitations; trajectory 4= prolonged hospitalization; trajectory 5= fatal.

TABLE 3 SUPPLEMENT

Demographics of survey respondents and non-respondents

|  |  | Non-respondents |  | Respondents |  | p-value |
| --- | --- | --- | --- | --- | --- | --- |
|  |  | Mean | % or SD^3^ (IQR) | Mean | % or SD^3^ (IQR) |  |
| Age |  | 58.7 | 16^3^ | 56.1 | 14.4^3^ | 0.12 |
| Sex | Male | 66 | 58.4 | 359 | 61.0 | 0.69 |
| Race^1^ | White | 50 | 44.2 | 289 | 49.1 | 0.38 |
|  | Black | 28 | 24.8 | 130 | 22.1 |  |
|  | Asian | 8 | 7.1 | 21 | 3.6 |  |
| Ethnicity^2^ | Non-Hispanic | 72 | 63.7 | 383 | 65.0 | 0.35 |
|  | Hispanic | 35 | 31.0 | 190 | 32.3 |  |
| Trajectory Group | 1 | 27 | 23.9 | 145 | 24.6 | 0.01 |
|  | 2 | 22 | 19.5 | 195 | 33.1 |  |
|  | 3 | 36 | 31.9 | 150 | 25.5 |  |
|  | 4 | 28 | 24.8 | 99 | 16.8 |  |
| Enrollment period | May 2020-September 2020 | 52 | 46 | 242 | 41.1 | 0.9999 |
|  | October 2020- March 2021 | 61 | 54 | 347 | 58.9 |  |

^1^ 22% other/declined/unknown/missing, ^2^ 3% not specified, ^3^SD: standard deviation

Trajectory 1= brief length of stay; trajectory 2= intermediate length of stay; trajectory 3= intermediate length of stay with discharge limitations; trajectory 4= prolonged hospitalization.

IQR: interquartile range

p- value from two-sample t-test for continuous age and chi-square test for all other categorical variables

FIGURE 1 SUPPLEMENT

a

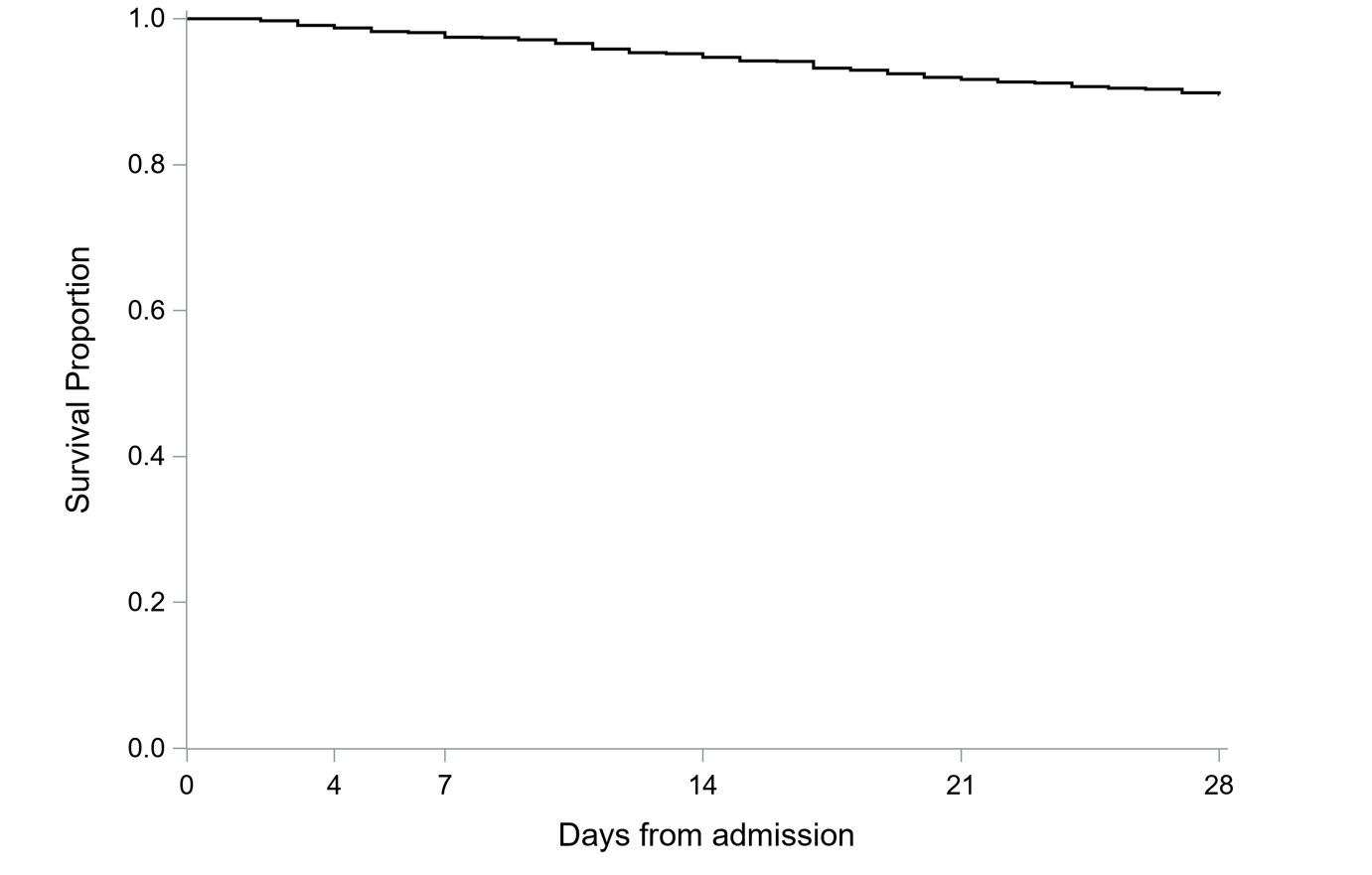

b

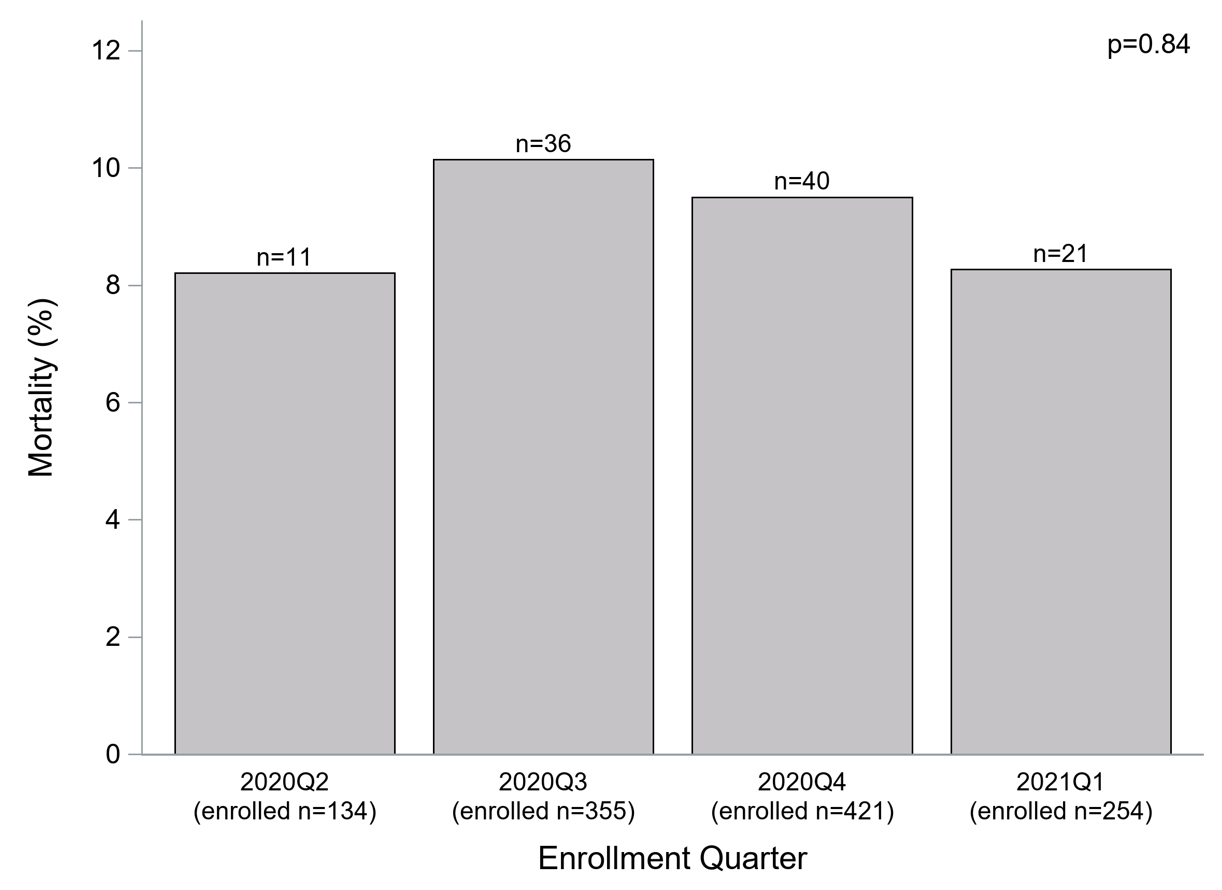

c

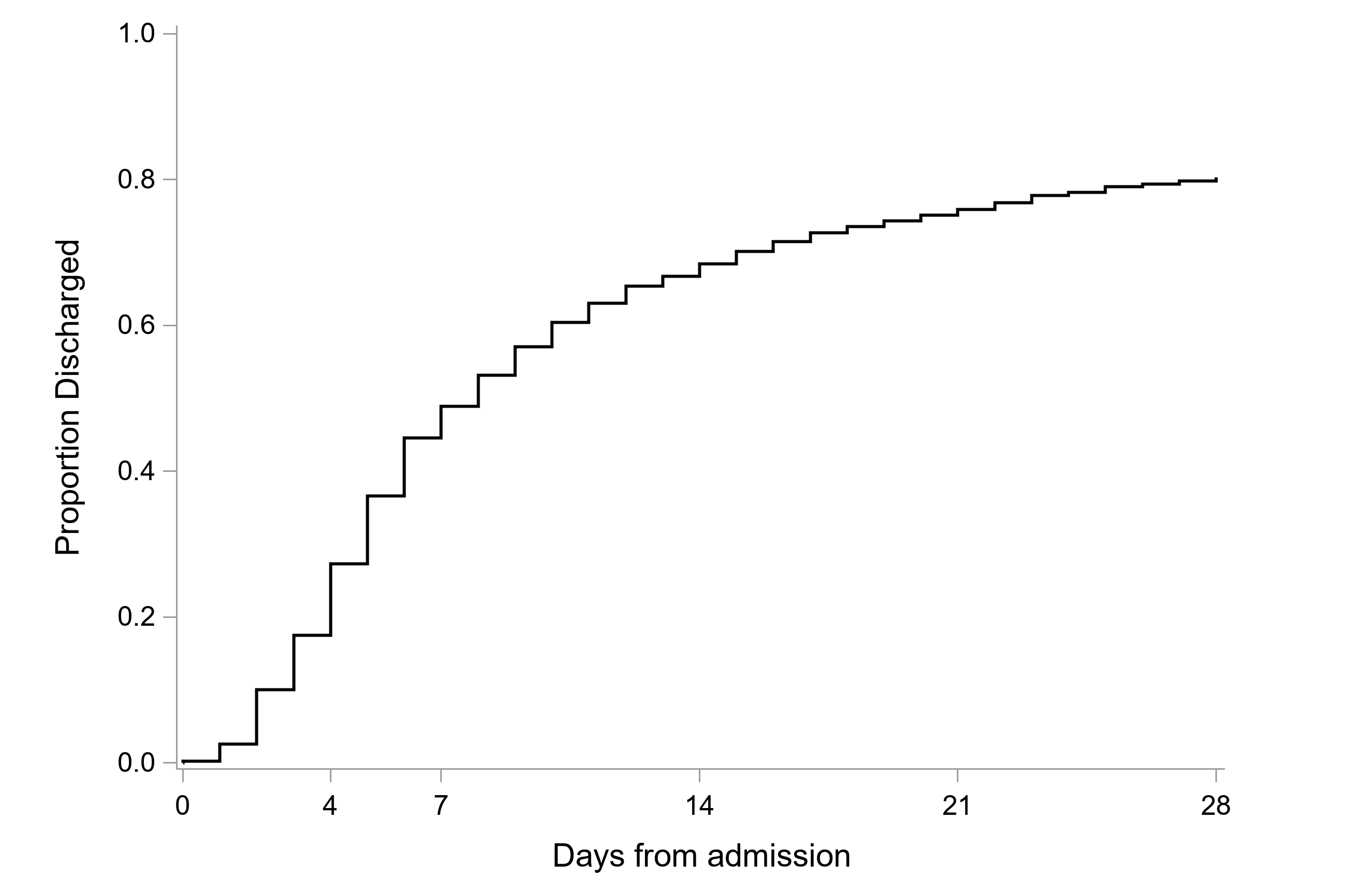

d
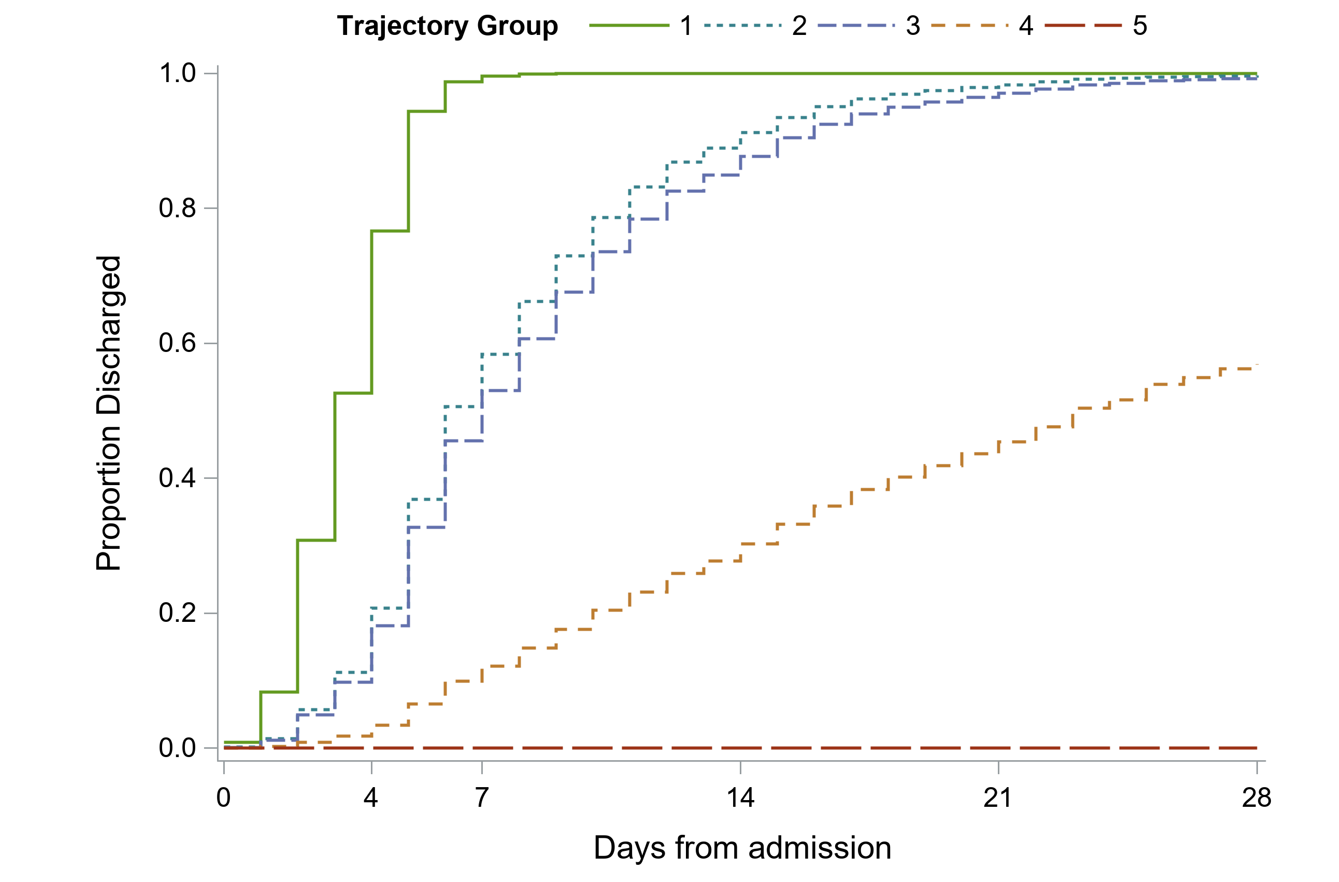

FIGURE 2 SUPPLEMENT

a

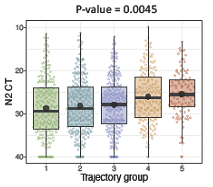

b

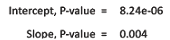

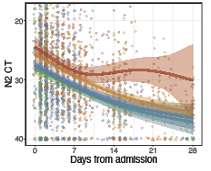

c

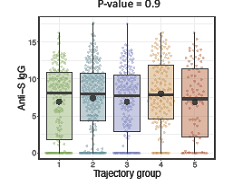

d

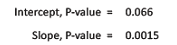

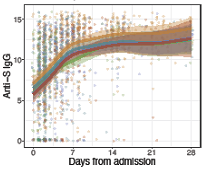

e

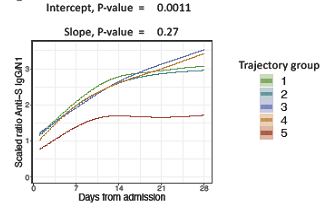

FIGURE 3 SUPPLEMENT

a

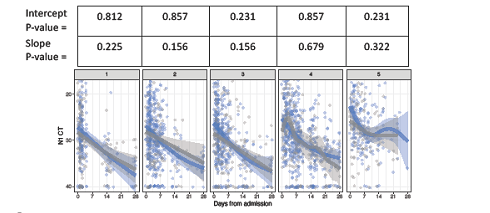

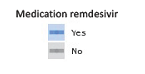

b

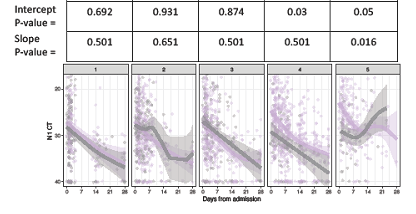

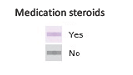

FIGURE 4 SUPPLEMENT

a

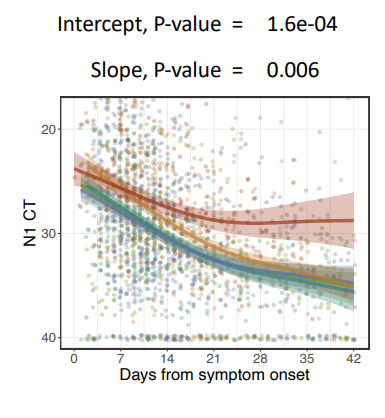

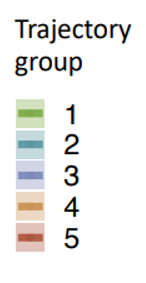

b

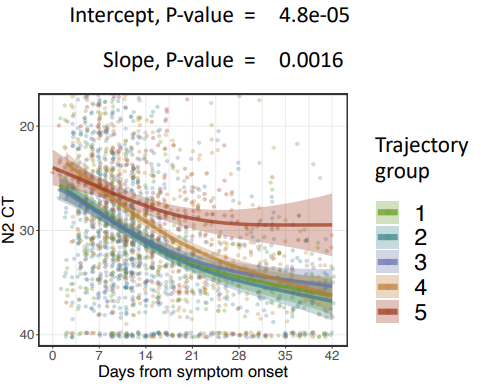

c

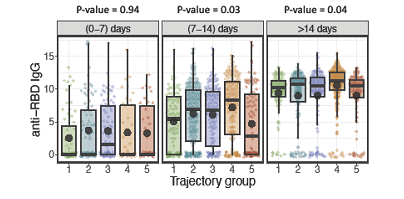

d

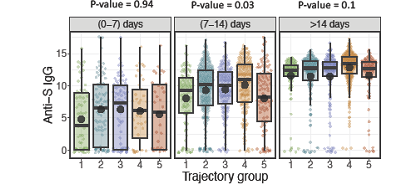

FIGURE 5 SUPPLEMENT

Identification of Pango lineages viral sequencing

a

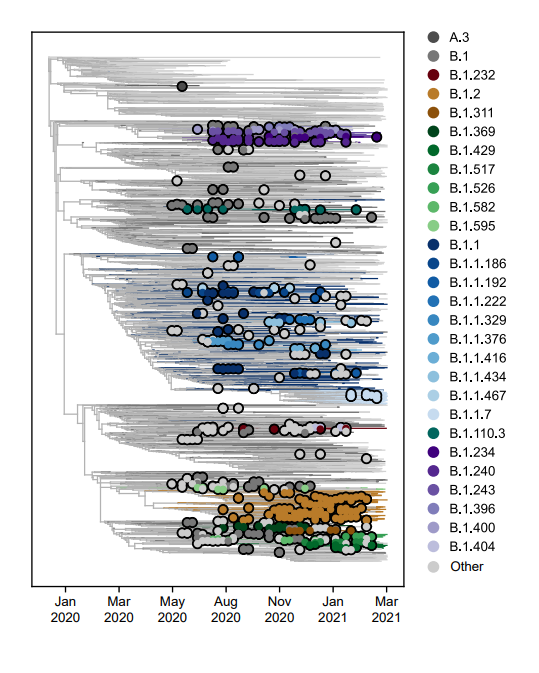

b

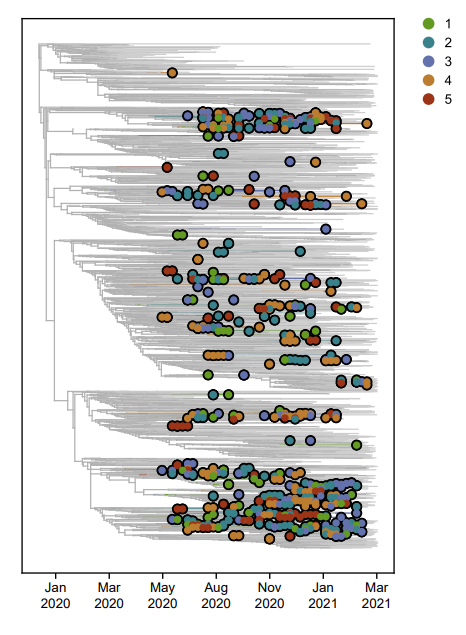

FIGURE 6 SUPPLEMENT

Frequency of symptoms reported at baseline and 3,6,9 and 12 months after discharge in survey respondents.

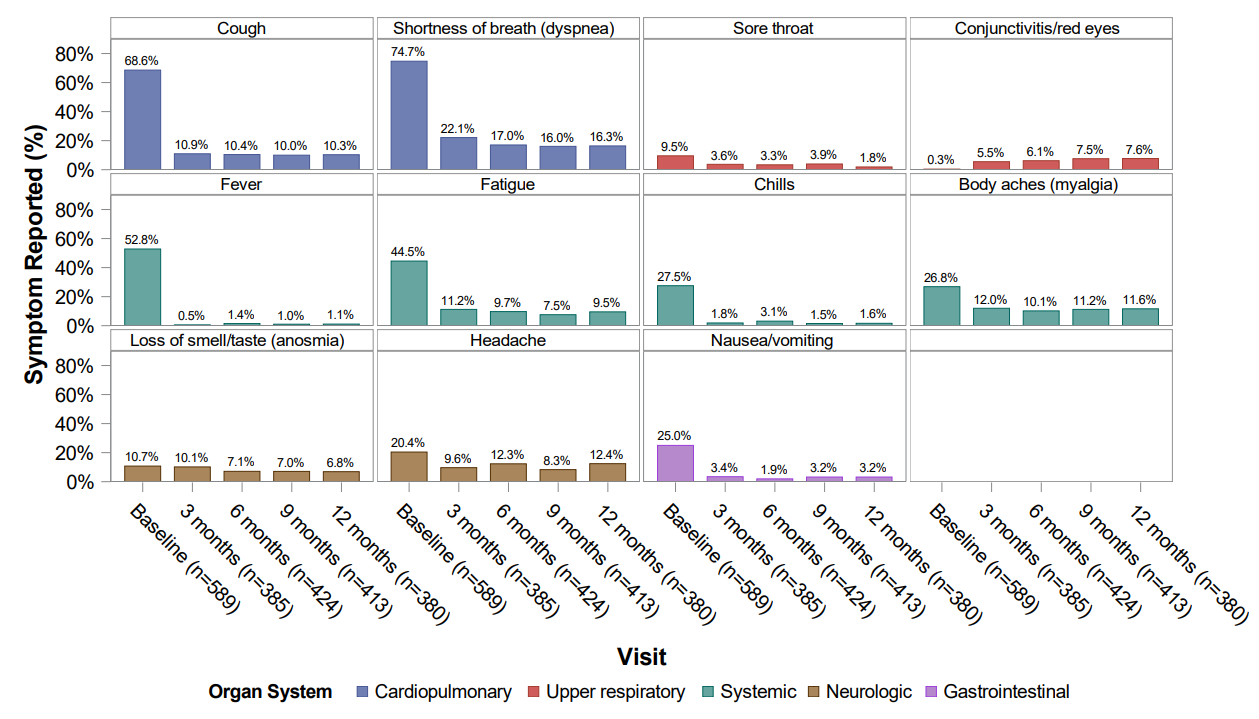

FIGURE 7 SUPPLEMENT

a

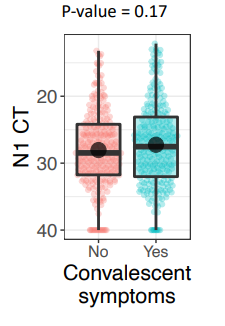

b

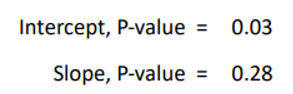

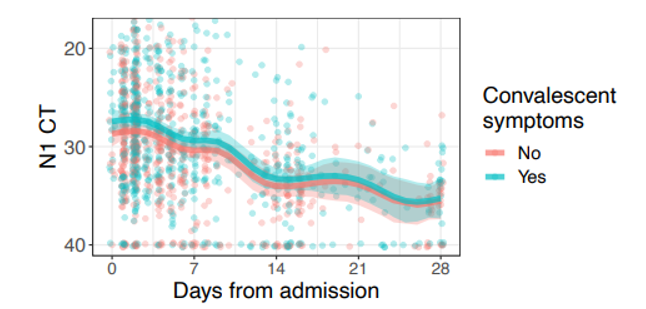

c

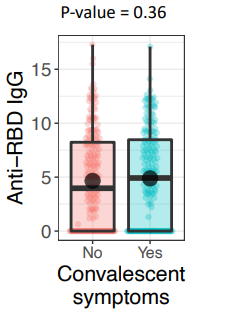

d

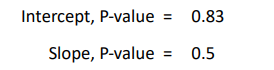

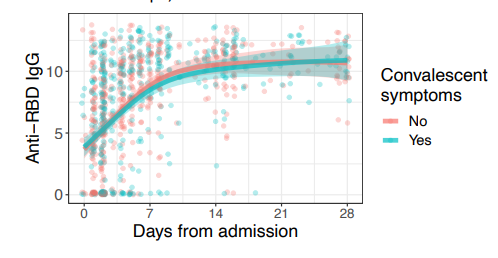

STROBE Statement—checklist of items that should be included in reports of observational studies

|  | Item No. | Recommendation | Page  No. | Relevant text from manuscript |
| --- | --- | --- | --- | --- |
| **Title and abstract** | 1 | (*a*) Indicate the study’s design with a commonly used term in the title or the abstract | 1 | Cohort |
|  |  | (*b*) Provide in the abstract an informative and balanced summary of what was done and what was found | 2 | Abstract Methods and Findings section |
| Introduction | | | |  |
| Background/rationale | 2 | Explain the scientific background and rationale for the investigation being reported | 7 | Introduction paragraph 1 |
| Objectives | 3 | State specific objectives, including any prespecified hypotheses | 9 | Outcomes Section |
| Methods | | | |  |
| Study design | 4 | Present key elements of study design early in the paper | 8-9 | Study design and setting section |
| Setting | 5 | Describe the setting, locations, and relevant dates, including periods of recruitment, exposure, follow-up, and data collection | 8-9 | Study design and setting section |
| Participants | 6 | (*a*) *Cohort study*—Give the eligibility criteria, and the sources and methods of selection of participants. Describe methods of follow-up  *Case-control study*—Give the eligibility criteria, and the sources and methods of case ascertainment and control selection. Give the rationale for the choice of cases and controls  *Cross-sectional study*—Give the eligibility criteria, and the sources and methods of selection of participants | Online supplement pages 1-2-3 | Online supplement study sections: Study participants – Data collection, study variables, and biologic samples. |
|  |  | (*b*) *Cohort study*—For matched studies, give matching criteria and number of exposed and unexposed  *Case-control study*—For matched studies, give matching criteria and the number of controls per case | N/A |  |
| Variables | 7 | Clearly define all outcomes, exposures, predictors, potential confounders, and effect modifiers. Give diagnostic criteria, if applicable | 9 | Outcomes Section |
| Data sources/ measurement | 8* | For each variable of interest, give sources of data and details of methods of assessment (measurement). Describe comparability of assessment methods if there is more than one group | eBIO online supplement DCF combined |  |
| Bias | 9 | Describe any efforts to address potential sources of bias | 11 | Statistics Section |
| Study size | 10 | Explain how the study size was arrived at | 38 | Figure 1: STROBE Cohort Diagram |

| Quantitative variables | 11 | Explain how quantitative variables were handled in the analyses. If applicable, describe which groupings were chosen and why | 11 | Statistics Section |
| --- | --- | --- | --- | --- |
| Statistical methods | 12 | (*a*) Describe all statistical methods, including those used to control for confounding | 11 | Statistics Section |
|  |  | (*b*) Describe any methods used to examine subgroups and interactions | 11 | Statistics Section |
|  |  | (*c*) Explain how missing data were addressed | 11 | Statistics Section |
|  |  | (*d*) *Cohort study*—If applicable, explain how loss to follow-up was addressed  *Case-control study*—If applicable, explain how matching of cases and controls was addressed  *Cross-sectional study*—If applicable, describe analytical methods taking account of sampling strategy | 21 | Caveats and Limitations |
|  |  | (*e*) Describe any sensitivity analyses | N/A |  |
| Results | | | | |
| Participants | 13* | (a) Report numbers of individuals at each stage of study—eg numbers potentially eligible, examined for eligibility, confirmed eligible, included in the study, completing follow-up, and analysed | 13 | Results section paragraph 1 |
|  |  | (b) Give reasons for non-participation at each stage |  |  |
|  |  | (c) Consider use of a flow diagram | 3 | Figure 1: STROBE Cohort Diagram |
| Descriptive data | 14* | (a) Give characteristics of study participants (eg demographic, clinical, social) and information on exposures and potential confounders | 13 | Results section paragraph 1 |
|  |  | (b) Indicate number of participants with missing data for each variable of interest | 37 | Figure 1: STROBE Cohort Diagram |
|  |  | (c) *Cohort study*—Summarise follow-up time (eg, average and total amount) | 9 | Data collection, study variables and biological samples |
| Outcome data | 15* | *Cohort study*—Report numbers of outcome events or summary measures over time | 38-46 | Figure 2 – Figure 3 |
|  |  | *Case-control study—*Report numbers in each exposure category, or summary measures of exposure |  |  |
|  |  | *Cross-sectional study—*Report numbers of outcome events or summary measures |  |  |
| Main results | 16 | (*a*) Give unadjusted estimates and, if applicable, confounder-adjusted estimates and their precision (eg, 95% confidence interval). Make clear which confounders were adjusted for and why they were included | 13-17 | Results section |
|  |  | (*b*) Report category boundaries when continuous variables were categorized | 13-17 | Results section |
|  |  | (*c*) If relevant, consider translating estimates of relative risk into absolute risk for a meaningful time period | N/A |  |

| Other analyses | 17 | Report other analyses done—eg analyses of subgroups and interactions, and sensitivity analyses | 47-51 | Figure 4- Table 1 |
| --- | --- | --- | --- | --- |
| Discussion | | | | |
| Key results | 18 | Summarise key results with reference to study objectives | 18 | Discussion paragraph 1 |
| Limitations | 19 | Discuss limitations of the study, taking into account sources of potential bias or imprecision. Discuss both direction and magnitude of any potential bias | 21 | Caveats and Limitations |
| Interpretation | 20 | Give a cautious overall interpretation of results considering objectives, limitations, multiplicity of analyses, results from similar studies, and other relevant evidence | 21-22 | Conclusion section |
| Generalisability | 21 | Discuss the generalisability (external validity) of the study results | 20-21 | Caveats and Limitations |
| Other information | |  | | |
| Funding | 22 | Give the source of funding and the role of the funders for the present study and, if applicable, for the original study on which the present article is based | 30-31 | Funding section |

*Give information separately for cases and controls in case-control studies and, if applicable, for exposed and unexposed groups in cohort and cross-sectional studies.

**Note:** An Explanation and Elaboration article discusses each checklist item and gives methodological background and published examples of transparent reporting. The STROBE checklist is best used in conjunction with this article (freely available on the Web sites of PLoS Medicine at http://www.plosmedicine.org/, Annals of Internal Medicine at http://www.annals.org/, and Epidemiology at http://www.epidem.com/). Information on the STROBE Initiative is available at www.strobe-statement.org.
