## Supplementary figures and images for "Phenotypes of disease severity in a cohort of hospitalized COVID-19 patients: results from the IMPACC study"

### Supplemental Figure 1

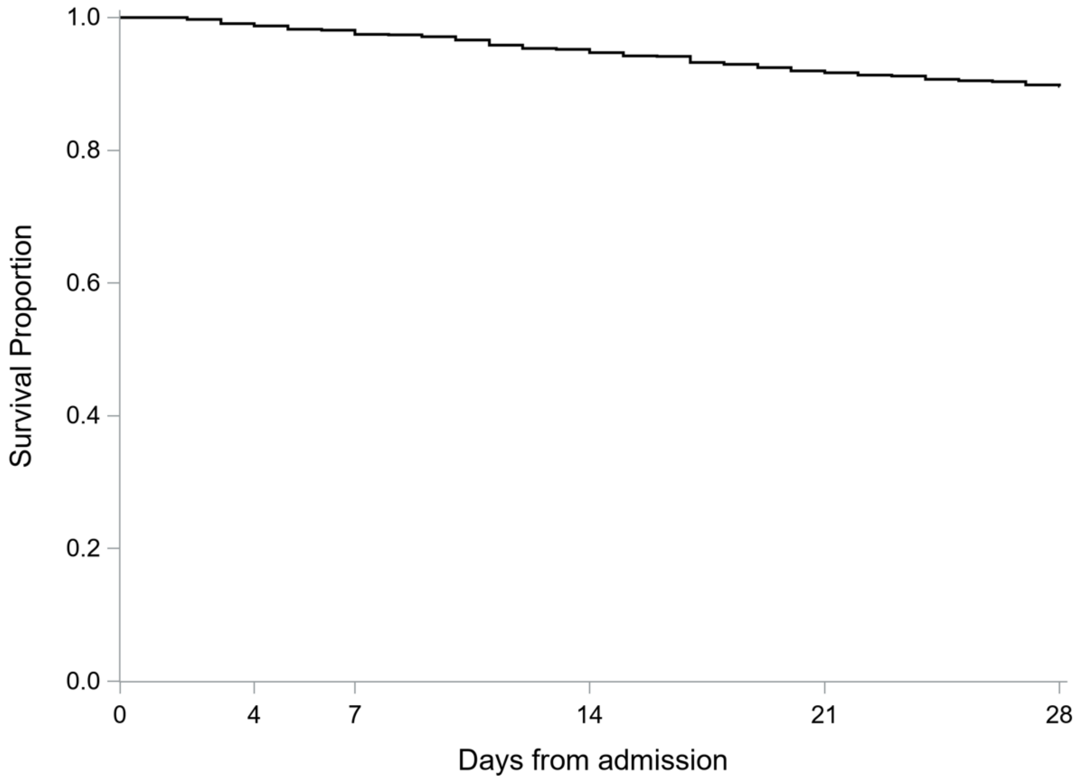

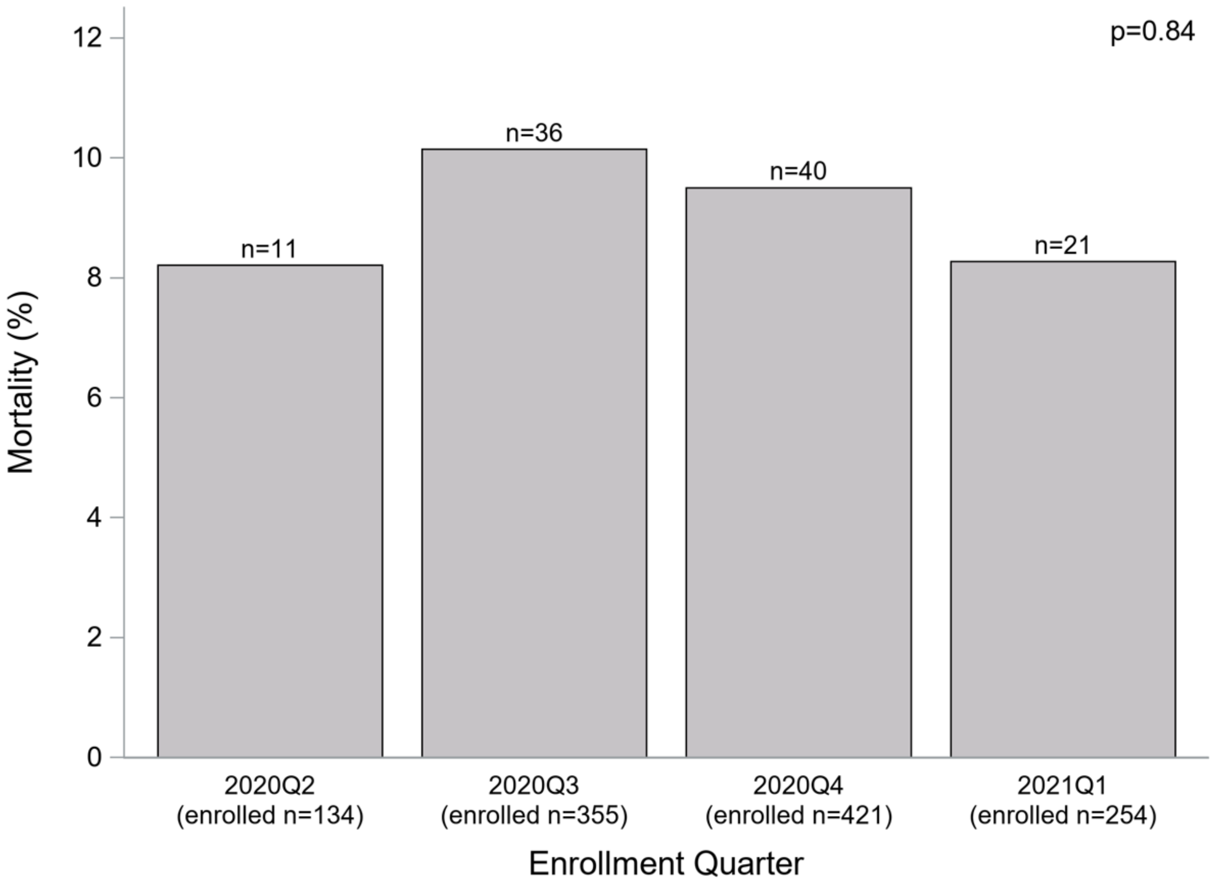

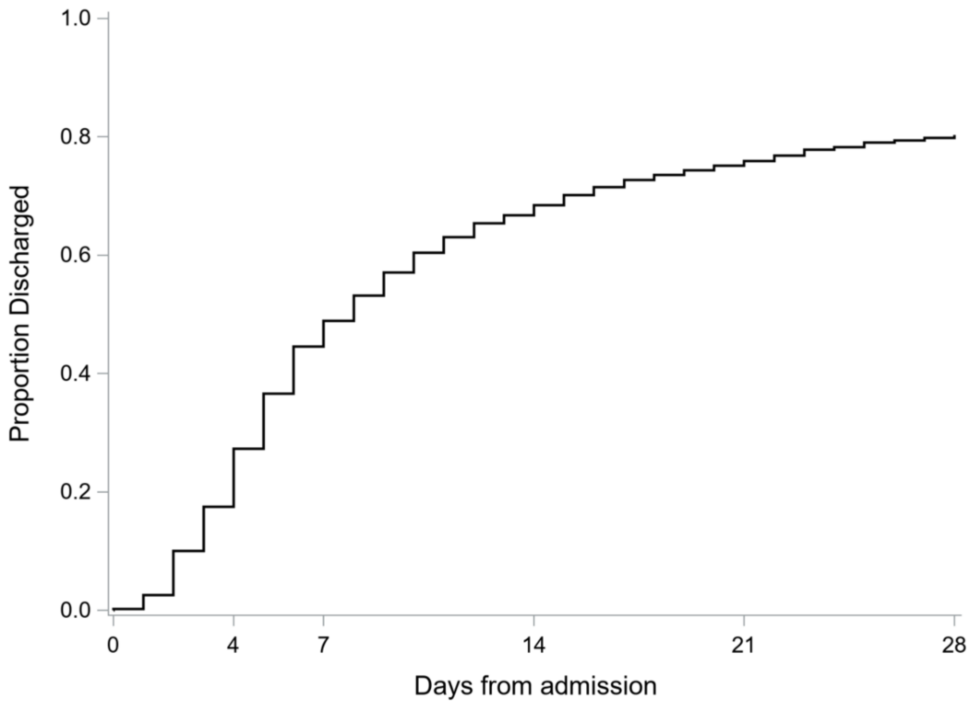

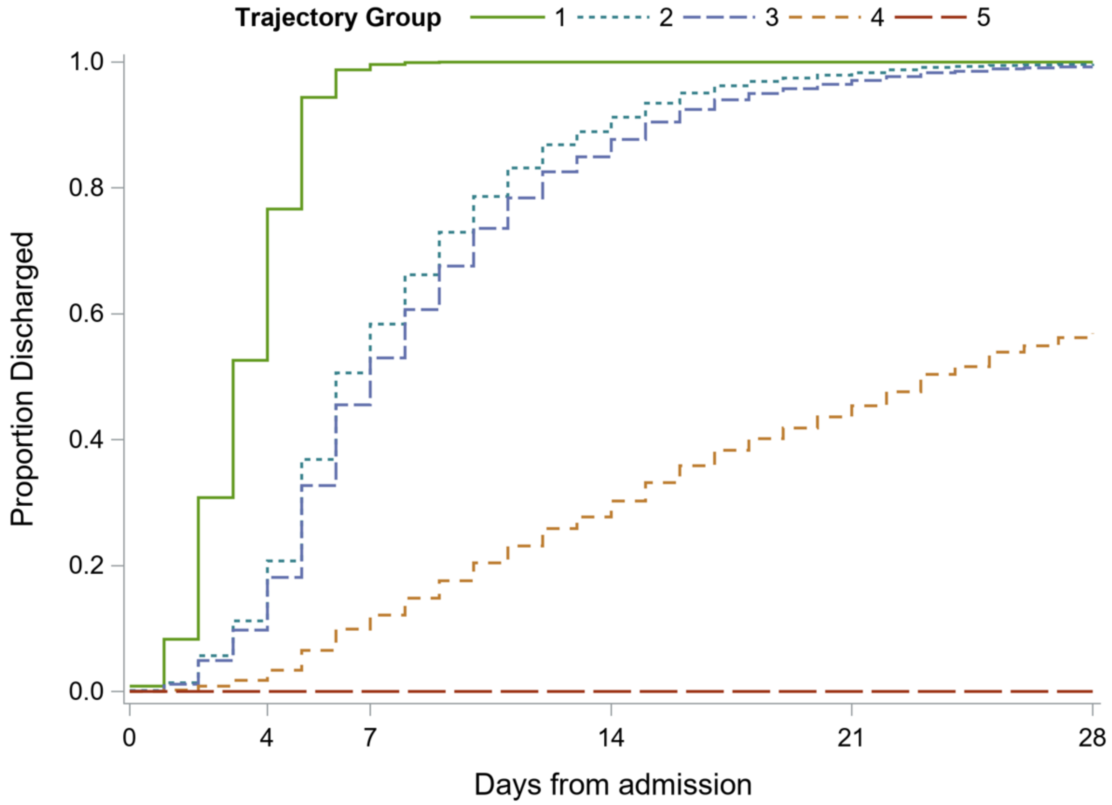

### Supplemental Figure 2

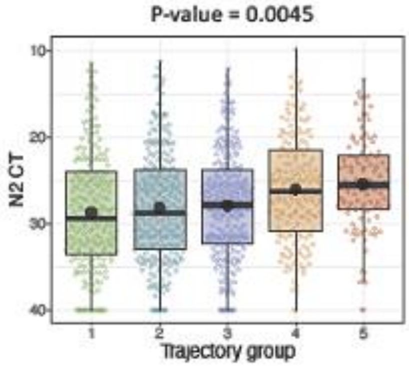

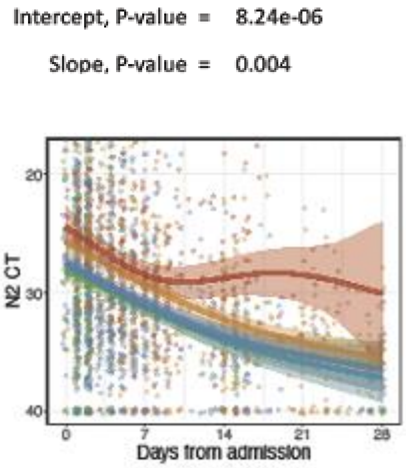

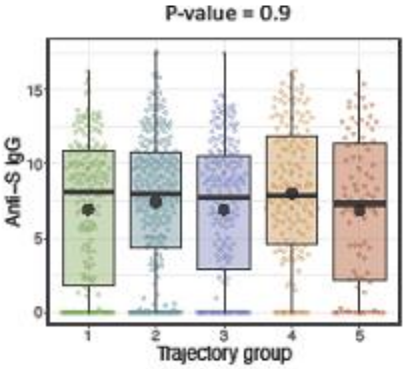

d

Intercept, P-value = 0.066

Slope, P-value = 0.0015

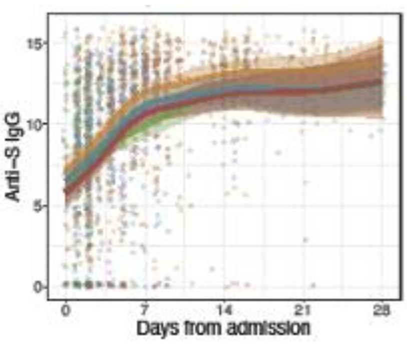


e

Intercept, P-value = 0.0011

Slope, P-value = 0.27

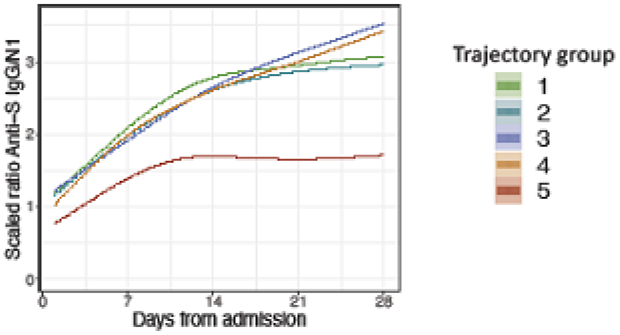

### Supplemental Figure 4

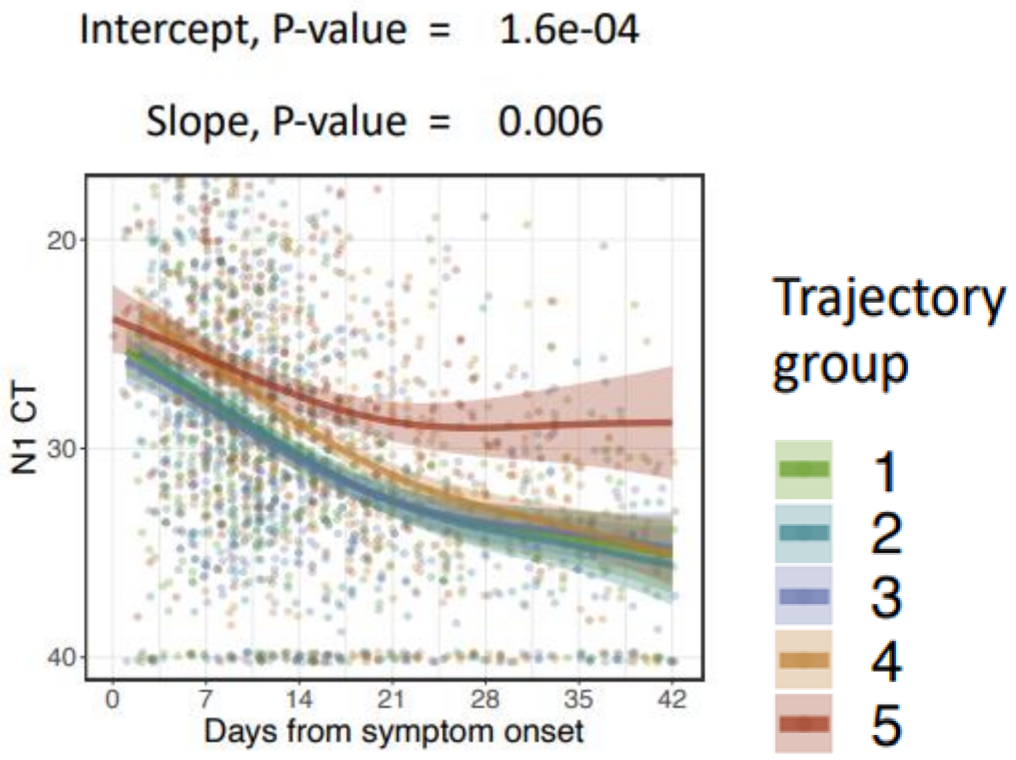

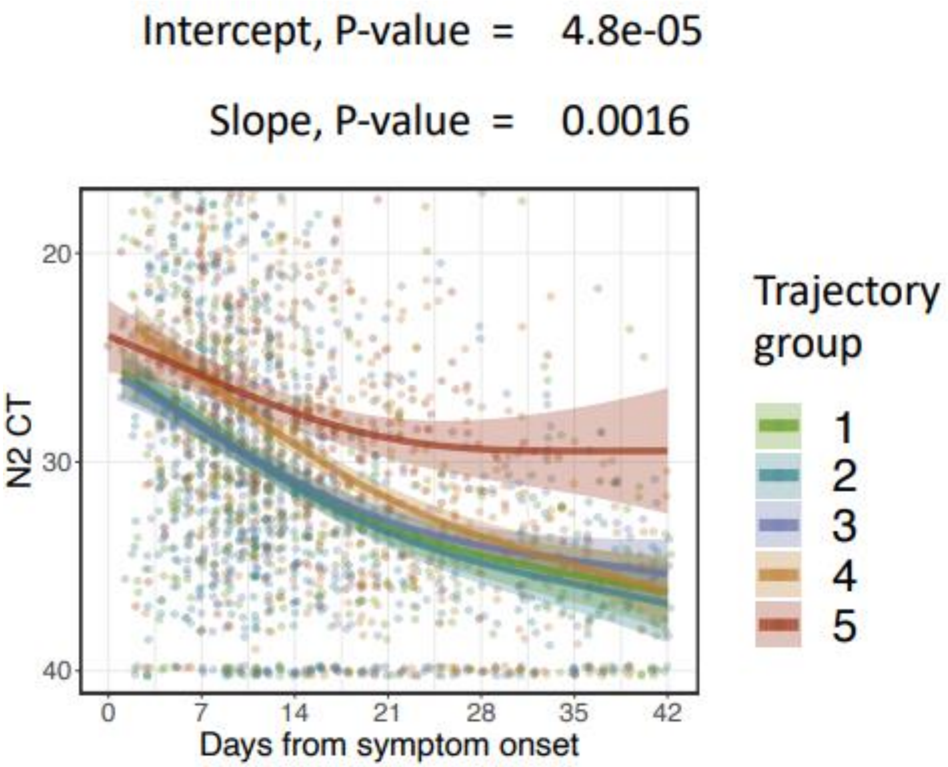

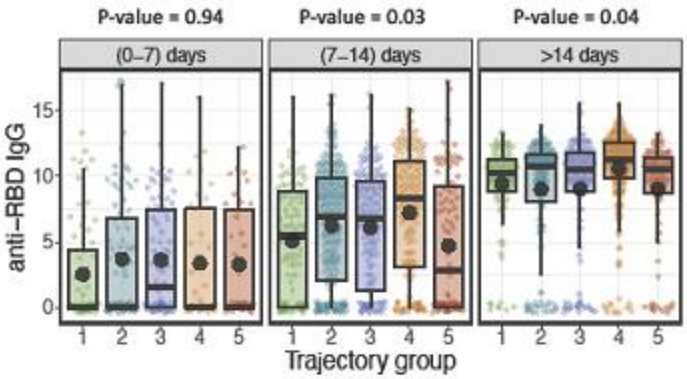

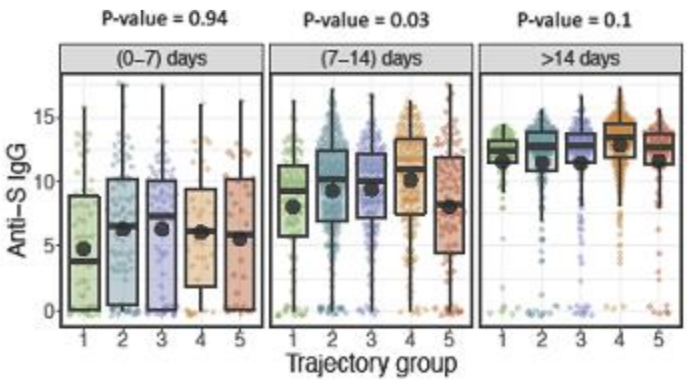

### Supplemental Figure 5

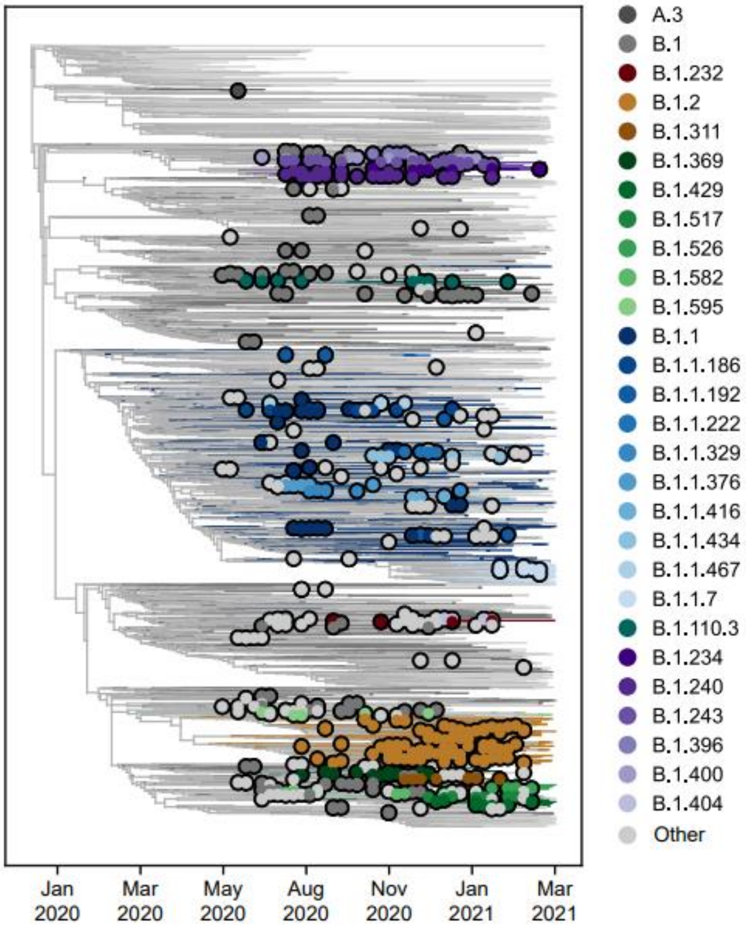

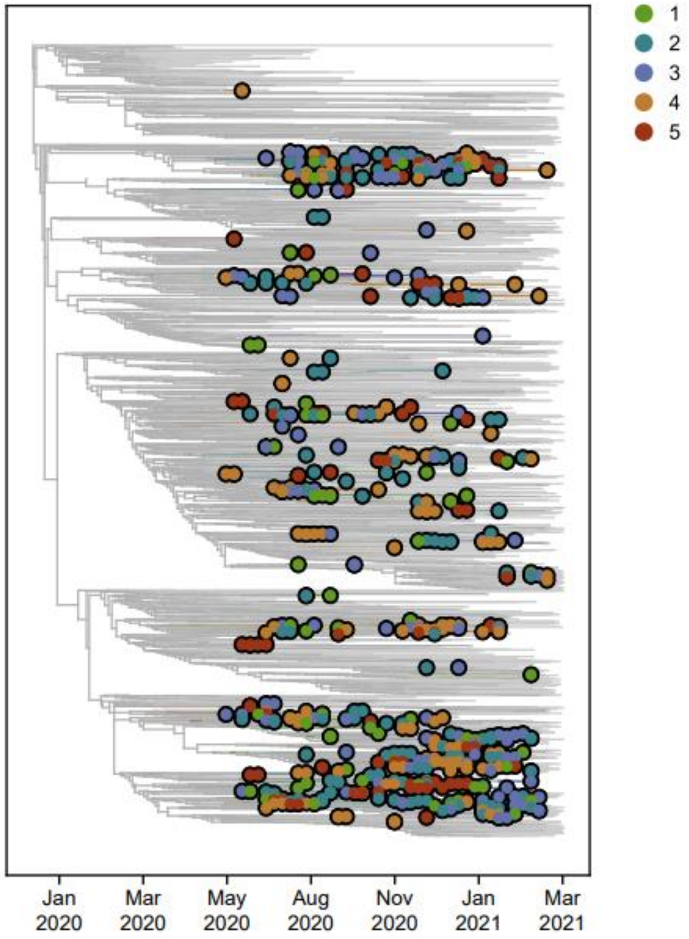

### Supplemental Figure 6

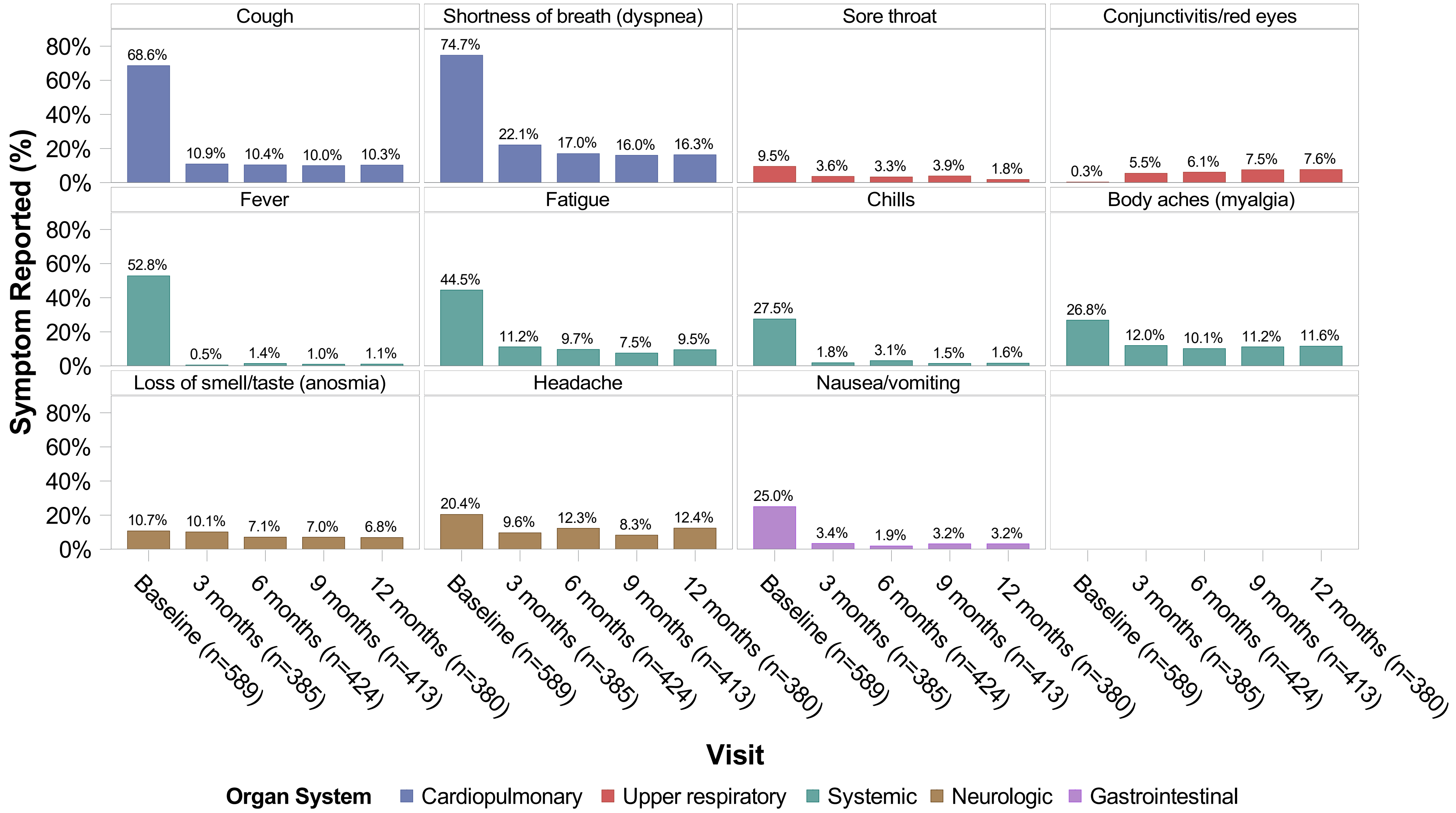

### Supplemental Figure 7

a

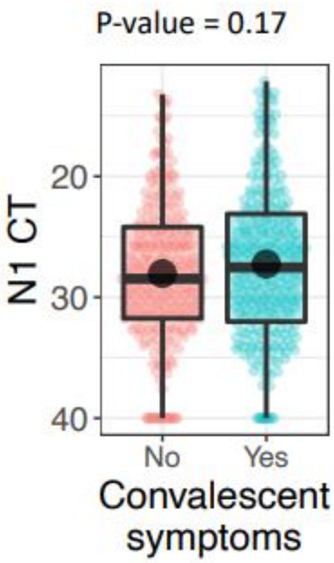

b

Intercept, P-value = 0.03

Slope, P-value = 0.28

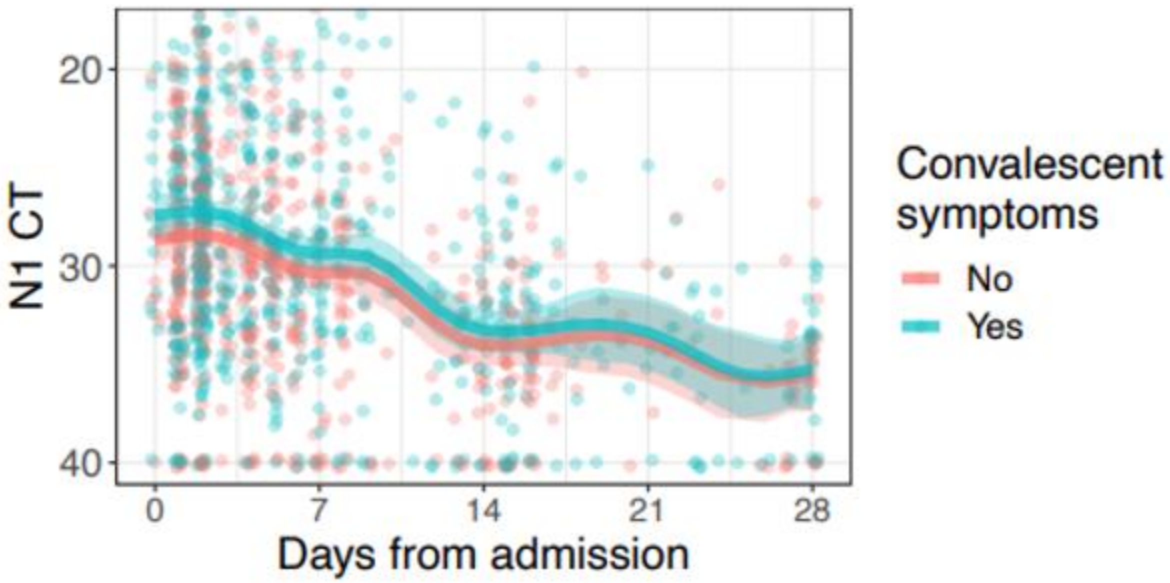

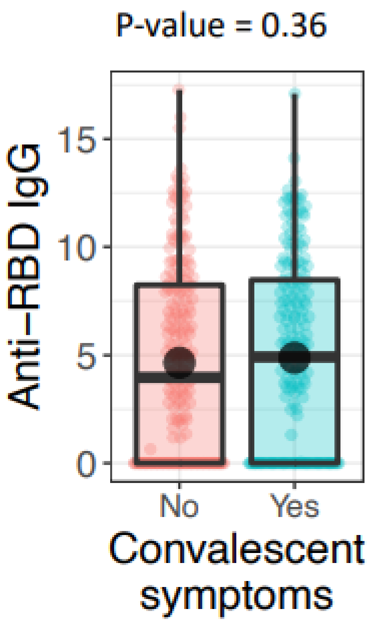

d

Intercept, P-value = 0.83

Slope, P-value = 0.5

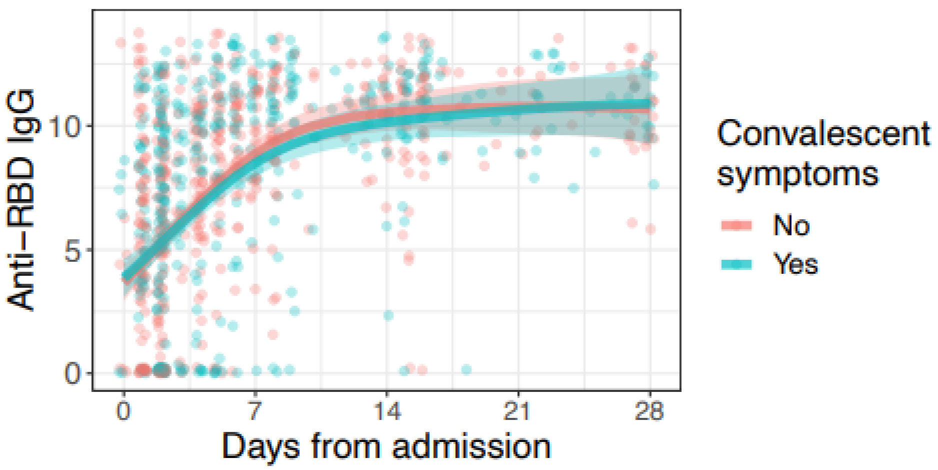
