## Supplemental Figure 3 for "Phenotypes of disease severity in a cohort of hospitalized COVID-19 patients: results from the IMPACC study"

a

| | | | | | |
|---|---|---|---|---|---|
| Intercept P-value = | 0.812 | 0.857 | 0.231 | 0.857 | 0.231 |
| Slope P-value = | 0.225 | 0.156 | 0.156 | 0.679 | 0.322 |

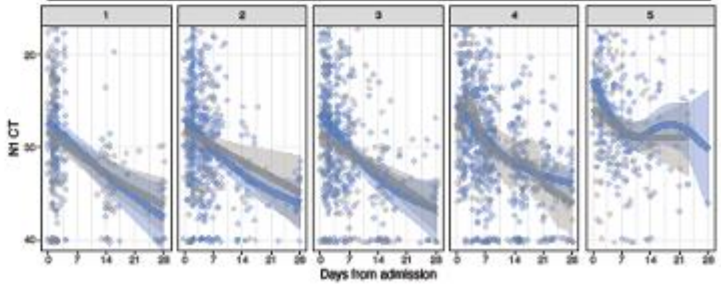


Medication remdesivir

Yes

No

b

| | | | | | |
|---|---|---|---|---|---|
| Intercept P-value = | 0.692 | 0.931 | 0.874 | 0.03 | 0.05 |
| Slope P-value = | 0.501 | 0.651 | 0.501 | 0.501 | 0.016 |

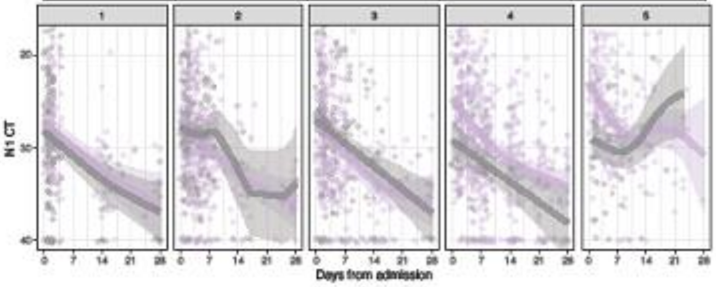


Medication steroids

- Yes
- No
